# Deep Learning-Based Multi-View Echocardiographic Framework for Comprehensive Diagnosis of Pericardial Disease

**DOI:** 10.1101/2025.07.24.25332108

**Authors:** Sihyeon Jeong, In Tae Moon, Jaeik Jeon, Dawun Jeong, Jina Lee, Jiyeon Kim, Seung-Ah Lee, Yeonggul Jang, Yeonyee E. Yoon, Hyuk-Jae Chang

**Author notes:** Corresponding authors: Yeonyee E. Yoon, MD, PhD, Professor, Division of Cardiology, Cardiovascular Center Seoul National University Bundang Hospital, 82, 173 Beon-gil, Gumi-ro, Bundang-gu, Seongnam-si, Gyeonggi-do, Republic of Korea, And, Yeonggul Jang, PhD, Ontact Health Inc., Seoul, Republic of Korea, 50-5, Ewhayeodae-gil, Seodaemun-gu, Seoul, Republic of Korea. The first two authors contributed equally to this work.

## Abstract

**Background:** Pericardial disease spans a wide spectrum from small effusions to life-threatening tamponade or constriction. Transthoracic echocardiography (TTE) is the main diagnostic tool, but its interpretation is limited by operator dependence and incomplete functional assessment. Existing deep learning (DL) models focus mainly on effusion detection, lacking broader evaluation.

**Methods:** We developed a DL-based framework that performs sequential assessment of pericardial disease: (1) morphological features, including effusion amount (normal/small/moderate/large) and pericardial thickening/adhesion (yes/no), from five B-mode views, and (2) hemodynamic significance (yes/no), incorporating Doppler and inferior vena cava measurements. The developmental dataset comprises 2,253 TTEs from multiple Korean institutions (225 for internal testing), and the independent external test set consists of 274 TTEs.

**Results:** In the internal test set, diagnostic accuracy was 81.8-97.3% for effusion, 91.6% for thickening/adhesion, and 86.2% for hemodynamic significance. External test set accuracy was 80.3-94.2%, 94.5%, and 85.5%, respectively. Area under the receiver operating curves (AUROCs) for the three tasks was 0.92-0.99, 0.90, and 0.79 internally, and 0.95-0.98, 0.85, and 0.76 externally. Sensitivity for thickening/adhesion and hemodynamic significance improved from 66.7% to 77.3%, and 68.8% to 80.8%, respectively, when poor image quality were excluded. Similar performance gains were observed in subgroups with complete target views and a higher number of available video clips.

**Conclusions:** This study presents the first DL-based TTE model for broader pericardial disease evaluation, integrating morphological with supportive functional assessments. The proposed framework demonstrated strong generalizability and aligned with the real-world diagnostic workflow. However, caution is warranted when interpreting results under suboptimal imaging conditions.

## 1. INTRODUCTION

Pericardial diseases are frequently encountered in clinical practice and encompass a wide clinical spectrum, ranging from small, asymptomatic effusions to severe and life-threatening conditions such as cardiac tamponade. While some cases resolve spontaneously without medical intervention, others may progress to constrictive pericarditis, characterised by the formation of pericardial adhesions and thickening. This clinical heterogeneity underscores the need for accurate and timely diagnostic approaches that integrate both structural and functional cardiac assessment^1,2^. Transthoracic echocardiography (TTE) is the primary non-invasive imaging modality for evaluating pericardial diseases, owing to its widespread availability, real-time imaging capabilities, and ability to assess cardiac hemodynamics^1^. However, the diagnostic performance of TTE remains highly dependent on operator expertise, particularly in image acquisition and interpretation^3^, which can induce variability and affect patient care. Tools that provide consistent and reproducible assessments may therefore offer meaningful support in daily clinical decision-making.

Recent advances in deep learning (DL) have shown promise in addressing these limitations of echocardiography. Nevertheless, most existing DL-based approaches have focused on isolated tasks, such as detecting pericardial effusion, without extending to the broader spectrum of pericardial disease. As a result, the clinical applicability of these models remains limited, particularly when timely identification of abnormalities is required to guide further expert evaluation. To address this gap and to improve applicability in real-world practice, the current study aimed to develop and validate a DL framework capable of identifying a broader spectrum of pericardial diseases using TTE. Rather than relying on a complete echocardiographic examination, the proposed model was designed to operate on a set of commonly available standard views, acknowledging that the availability and completeness of imaging can vary in real-world clinical practice. The framework performs a two-stage evaluation: (1) identification of morphological features, including effusion, thickening, and adhesion, and (2) supportive assessment of hemodynamic significance using automatically extracted Doppler and inferior vena cava (IVC) parameters when available. To ensure generalizability, the framework was validated not only on internal test data but also on an independent external dataset. In addition, as a secondary exploratory aim, we evaluated the impact of image quality (IQ) and data completeness (availability of target views and number of clips) on model performance, acknowledging these factors as inherent challenges in real-world echocardiographic practice.

## 2. METHODS

### 2.1. Study Population

The DL-based framework developed in this study was trained and validated using the Open AI Dataset Project (AI-Hub) dataset, a national echocardiographic database supported by the Ministry of Science and ICT of the Republic of Korea^4–7^. This dataset comprises approximately 30,000 TTE examinations retrospectively collected from multiple tertiary hospitals in South Korea between 2012 and 2021, encompassing a broad spectrum of cardiovascular conditions. For model development, 2,253 TTE examinations were selected from 2,115 patients, including both individuals with pericardial diseases and normal controls (**Supplemental Methods 1**). Normal cases were drawn from the “normal” category and consisted of asymptomatic individuals who underwent TTE examinations during routine health check-ups, with only those showing normal pericardial findings included. Pericardial disease cases were extracted from the “pericardial disease” category, identified using ICD-10 diagnostic codes (I30.9, I31.1, I31.3, I31.9, and C79.88). Then the diagnostic categories were determined based on the presence or absence of pericardial effusion, pericardial thickening and adhesions, and hemodynamic significance, as described in **Supplemental Methods 2**. The number of cases corresponding to each diagnostic category was as follows: normal pericardium (n=1,178), pericardial effusion without complication (n=703), cardiac tamponade (n=138), pericarditis with effusion (n=35), effusive-constrictive pericarditis (n=98), pericarditis without constriction (n=14), and constrictive pericarditis (n=87). The developmental dataset was randomly divided into training, validation, and internal test sets in an 8:1:1 ratio, comprising 1,665, 225, and 225 patients, respectively (corresponding to 1,803, 225, and 225 TTE examinations) (**Table 1**). When a patient had multiple TTE examinations, all examinations from that patient were assigned to the training set to prevent data leakage. Diagnostic labels were assigned to the examination label, as pericardial status may vary over time, even within the same individual.

**Table 1.** Baseline Characteristics.

|  | Developmental Dataset |  |  | p-value * | External Test | p-value † |
| --- | --- | --- | --- | --- | --- | --- |
|  | Training | Validation | Internal Test |  |  |  |
| **No. of Patients** | **1665** | **225** | **225** |  | **255** |  |
| Age, years | 59.0 ± 17.9 | 58.4 ± 16.8 | 60.0 ± 17.8 | 0.619 | 67.9 ± 16.9 | <0.001 |
| Male, n (%) | 807 (48.5%) | 117 (52.0%) | 98 (43.6%) | 0.514 | 122 (47.8%) | 0.365 |
| BMI, kg/m<sup>2</sup> | 23.6 ± 3.9 | 23.9 ± 4.8 | 23.5 ± 4.1 | 0.177 | 23.9 ± 4.0 | 0.298 |
| **No. of TTE exams** | **1803** | **225** | **225** |  | **274** |  |
| Pericardial effusion, n (%) | 762 | 107 | 104 |  | 198 |  |
| Small | 252 (14.0%) | 35 (15.6%) | 28 (12.4%) | 0.636 | 100 (36.5%) | <0.001 |
| Moderate | 312 (17.3%) | 47 (20.9%) | 54 (24.0%) | 0.029 | 54 (19.7%) | 0.294 |
| Large | 198 (11.0%) | 25 (11.1%) | 22 (10.0%) | 0.855 | 44 (16.1%) | 0.054 |
| Pericardial thickness or adhesion, n (%) | 176 (9.8%) | 22 (9.0%) | 33 (14.7%) | 0.109 | 19 (6.9%) | 0.008 |
| Hemodynamic significance, n (%) | 261 (14.5%) | 28 (12.4%) | 34 (15.1%) | 0.672 | 32 (11.7%) | 0.321 |
| **Pericardial Disease Category, n (%)** |  |  |  |  |  |  |
| Normal pericardium | 961 (53.3%) | 110 (48.9%) | 107 (47.6%) | 0.149 | 65 (23.7%) | < 0.001 |
| Pericardial effusion without complication | 552 (30.6%) | 77 (34.2%) | 74 (32.9%) | 0.462 | 160 (58.4%) | <0.001 |
| Cardiac tamponade | 114 (6.3%) | 13 (5.8%) | 11 (4.9%) | 0.681 | 30 (11.0%) | 0.022 |
| Pericarditis with effusion | 23 (1.3%) | 7 (3.1%) | 5 (2.2%) | 0.077 | 8 (2.9%) | 0.838 |
| Effusive-constrictive pericarditis | 73 (4.0%) | 10 (4.4%) | 15 (6.7%) | 0.192 | 0 (0.0%) | < 0.001 |
| Pericarditis without constriction | 6 (0.3%) | 3 (1.3%) | 5 (2.2%) | 0.001 | 9 (2.9%) | 0.658 |
| Constrictive pericarditis | 74 (4.1%) | 5 (2.2%) | 8 (3.6%) | 0.373 | 2 (0.07%) | 0.049 |

\*p-values indicate comparisons among the training, validation, and internal test sets.
†p-value represent comparisons between the internal test set and the external test set.

In addition, an external test set was constructed using 274 TTE examinations from 255 patients collected at Uijeongbu Eulji University Hospital between 2021 and 2024. For this dataset, normal controls were randomly selected from individuals whose TTE examinations were reviewed and confirmed to show normal pericardial findings. Pericardial disease cases were also collected to ensure broad representation of diverse disease phenotypes. The diagnostic categories in the external test set were as follows: normal pericardium (n=65), pericardial effusion without complication (n=160), cardiac tamponade (n=30), pericarditis with effusion (n=8), pericarditis without constriction (n=9), and constrictive pericarditis (n=2). The study followed the Declaration of Helsinki (as revised in 2013). The institutional review board of each hospital approved this study and waived the requirement for informed consent because of the retrospective design and observational nature of the study (2021-0147-003/CNUH 2021-04-032/ HYUH 2021-03-026-003/SCHBC 2021-03-007-001/B 2104/677-004). Additionally, to establish and utilise the external test dataset, further IRB approval was obtained from Uijeongbu Eulji University Hospital (2025-03-018). All clinical and echocardiographic data were fully anonymised before data analysis.

### 2.2. TTE Acquisition and Utilisation

All TTE examinations were performed by trained echocardiographers or cardiologists and initially interpreted by board-certified cardiologists specialising in echocardiography, under current clinical guidelines^1,8^. To improve labelling accuracy, two experienced cardiologists (S.A. Lee, with over 10 years of experience; YE Yoon, with over 15 years of experience) independently reviewed the entire TTE examinations for each case and reached a consensus on key diagnostic features. These included (1) the presence and amount of pericardial effusion (categorised as none, small, moderate, or large), (2) the presence of pericardial thickening and/or adhesion (yes or no), and (3) the presence of hemodynamic significance (yes or no). For the external test set, to reflect real-world practice, key diagnostic features were primarily determined based on the original TTE reports from the contributing institution. This labelling process was independently reviewed and confirmed by an experienced cardiologist (I.T. Moon, with over 7 years of experience), who was not involved in labelling the developmental dataset.

Importantly, while expert annotations were based on a comprehensive review of the full TTE examination, only a subset of standard views was provided as input to the DL framework. These selected views included parasternal long-axis (PLAX), parasternal short-axis (PSAX), apical four-chamber (A4C), A4C right ventricular (RV)-focused or modified views, and subcostal four-chamber (SC4C) views. Additionally, mitral inflow pulsed-wave (PW) Doppler, septal tissue Doppler imaging (TDI), and subcostal long-axis IVC views were included for automated measurement and incorporated into model input.

All TTE examinations were included in the developmental and external test sets, regardless of image completeness or quality, to reflect real-world clinical conditions. Cases were not excluded based on the absence of specific target views or suboptimal IQ. When multiple video clips were available for a given view, all available data were utilised as input for the DL-based model. Conversely, the absence of a particular view did not lead to the exclusion of the case. We additionally evaluated the effect of IQ and data completeness on model performance within the internal test set. For IQ assessment, the overall IQ of each case in the internal test set was independently assessed by I.T. Moon, based on the internally defined criteria (**Supplementary Methods 3**). IQ was classified as good, fair, or poor based on the clarity of pericardial structures and the presence of artefacts affecting morphologic or hemodynamic assessment. Data completeness was defined according to (1) availability of all five target B-mode views and (2) number of available B-mode video clips, stratified into tertiles.

### 2.3. DL-Based Framework

We propose a two-stage DL-based framework that sequentially models morphological and hemodynamic features of pericardial disease using multi-view echocardiographic data (**Figure 1**). This design reflects the clinical diagnostic workflow, which begins with a structural assessment and is followed by a functional evaluation. To obtain the required inputs, we used our previously developed artificial intelligence (AI)-based echocardiographic automatic view classification system (Sonix Health; Ontact Health Co., Ltd, Korea)^5–7,9,10^. All selected views were then reviewed by investigators to exclude misclassified clips and to retrieve missing views, ensuring consistent and accurate inputs for the model.

**Figure 1.**
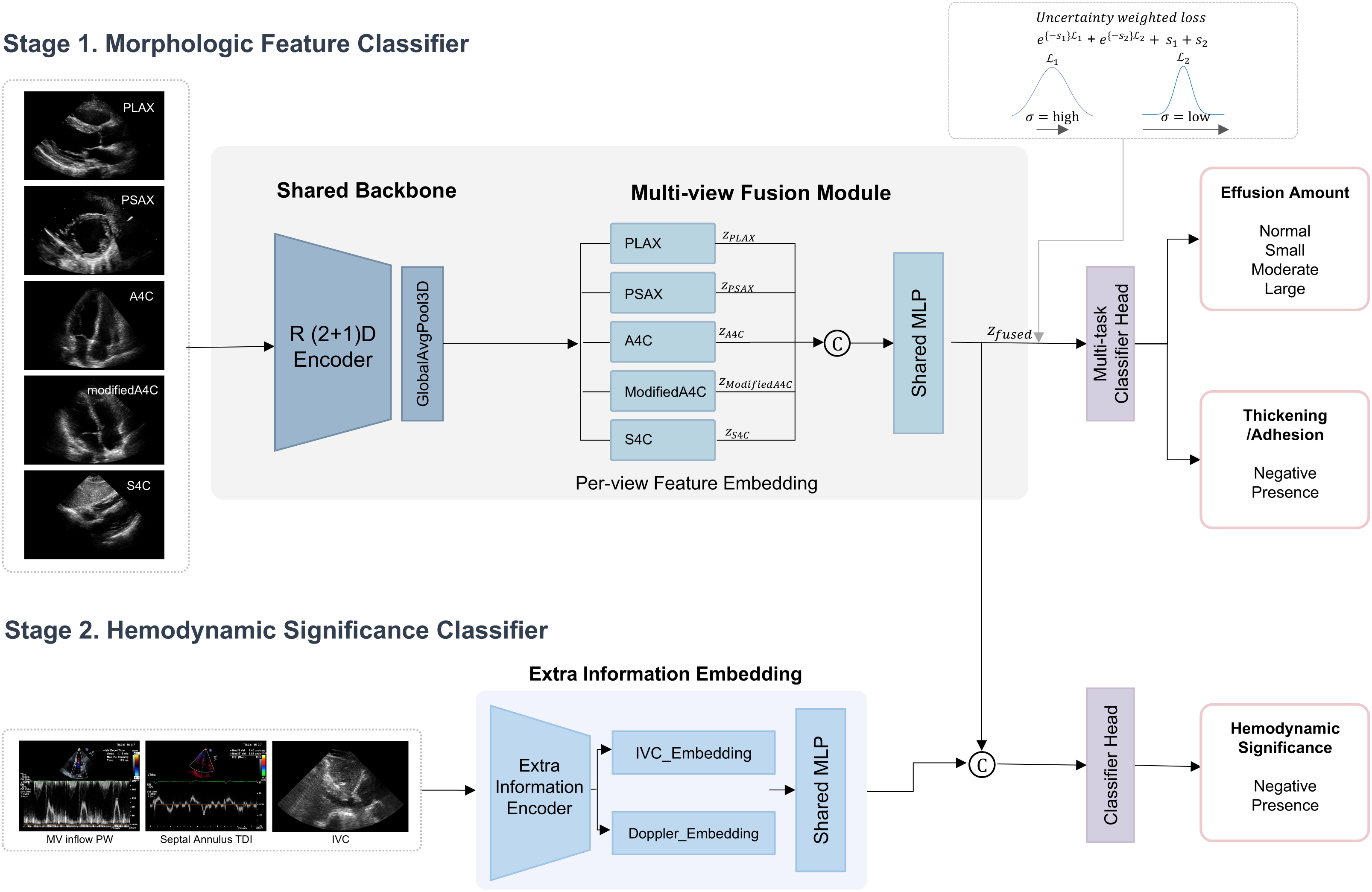
Two-Stage Deep Learning Framework for Pericardial Disease Assessment. The framework sequentially evaluates morphological and hemodynamic features, reflecting the clinical diagnostic workflow. **(A) Stage 1: Morphologic Feature Classifier** - Multi-view B-mode echocardiographic videos (PLAX, PSAX, A4C, Modified A4C, and S4C) are processed by a shared R(2+1)D encoder. Per-view spatiotemporal features are fused and used for multi-task classification of pericardial effusion and thickening/adhesion. An uncertainty-weighted loss accounts for task imbalance. **(B) Stage 2: Hemodynamic Significance Classifier** - Functional features from Doppler and IVC analysis are encoded and combined with morphological embeddings to predict hemodynamic significance.

In the first stage, the model processes B-mode echocardiographic video clips from selected views, including PLAX, PSAX, A4C, A4C RV-focused/modified, and SC4C, to extract morphological features indicative of pericardial effusion and pericardial thickening or adhesion. By leveraging multiple complementary views, the model addresses the limitations of single-view analysis and better captures the three-dimensional characteristics of the pericardial space. A modified R(2+1)D-18 network^11,12^ was used to extract spatiotemporal features from multi-view echocardiographic videos, and multi-view representations were aggregated through global pooling and concatenation. Detailed training procedures and preprocessing steps are described in **Supplemental Methods 4**. When multiple video clips were available for a given view, we explored several inference strategies to handle redundant inputs. Based on this comparison, we adopted an exhaustive combination-based probability aggregation method, which evaluated all possible multi-view and multi-clip combinations and averaged their predicted probabilities to improve robustness. Details of the compared methods and their results are provided in **Supplemental Methods 5.**

In the second stage, the model evaluates the hemodynamic significance of the morphological findings by incorporating additional functional signals from Doppler and IVC analysis. Both Doppler and IVC measurements were automatically extracted using our in-house AI-based models (SONIX Health, Ontact Health Co., Ltd, Korea), with the IVC analysis model being newly developed and introduced in this study. Doppler parameters, including mitral inflow PW Doppler (E, A, deceleration time), and septal annulus TDI (S’, E’, A’) were automatically measured^7,13^. When multiple spectral Doppler images were available, the image with the clearest Doppler signal was selected, and all cycles within the selected image were analysed to compute the mean input values. Respiratory variation of mitral or tricuspid inflow and simultaneous septal-lateral annulus TDI evaluation were not incorporated, as such acquisitions were rarely available and could bias the model toward operator suspicion. For IVC analysis, the longest full-length subcostal long-axis IVC video was selected to ensure reliable respiratory variation assessment. IVC dilatation and respiratory variation were quantified by our AI-based IVC segmentation model, which measured IVC diameter using heatmap-guided region extraction and segmentation area analysis (**Supplementary Methods 6**). Full-length subcostal long-axis IVC videos were used to ensure accurate automatic measurement. Based on automatic measurements, IVC dilatation was defined as a maximum diameter ≥21mm, and IVC plethora was defined as a lack of inspiratory collapse ≥50%. IVC dilatation status and plethora status were provided as model inputs. All functional features were integrated with morphological embeddings to support the detection of the hemodynamic significance of pericardial disease.

### 2.4. Validation and Statistical Analysis

The performance of each stage in our DL-based framework was validated using both internal and external test sets. Diagnostic performance was assessed using the area under the receiver operating characteristic curve (AUROC), accuracy, precision, sensitivity, specificity, and F1-score. Sampling variability was quantified by exhaustively evaluating every multi-view and multi-clip combination for each exam in the internal (n = 225) and external (n = 274) cohorts, averaging the resulting probabilities to yield a single deterministic prediction per exam (**Supplemental Methods 5**); this approach obviated the need for bootstrap resampling. Model inference was performed on each resampled set, and diagnostic metrics were recalculated.

For the prediction of pericardial thickening/adhesion and hemodynamic significance, the optimal probability cutoff was determined from the internal validation set using Youden’s J statistic. This cutoff was fixed and consistently applied in all model predictions, including bootstrap replicates. Additionally, the model’s performance in detecting hemodynamic significance was evaluated in a stepwise manner using three different input configurations. First, predictions were generated using only the five B-mode TTE videos employed in the initial stage. Second, performance was assessed by adding Doppler measurement, including mitral inflow PW Doppler and septal TDI. Third, the model incorporated additional IVC measurements. For each configuration, diagnostic performance was evaluated using confusion matrices, diagnostic metrics, and AUROC values.

## 3. RESULTS

### 3.1. Baseline Characteristics

Baseline characteristics and the distribution of pericardial disease across datasets are shown in **Table 1**. At the patient level, no significant differences were observed among the training, validation, and internal test sets in terms of age, sex, and body mass index (BMI). However, patients in the external test set (mean age 67.9 ± 16.9 years) were older than those in the internal test set (60.0 ± 17.8 years, p<0.001). Based on TTE examinations, compared to the internal test set, the external test set included more cases of small pericardial effusion (36.5% versus 12.4%, p<0.001), and fewer cases of pericardial thickening or adhesion (6.9% versus 14.7%, p = 0.008), with no significant difference in the presence of hemodynamic significance (11.7% versus 15.1%, p=0.321). The external test examinations also showed a lower proportion of normal pericardium (23.7% versus 47.6%, p<0.001), a higher proportion of uncomplicated pericardial effusion (58.4% versus 32.9%, p<0.001), and more frequent cardiac tamponade (11.0% versus 4.9%, p=0.022). Effusive-constrictive pericarditis was not observed in the external cohort, while other disease categories were similarly distributed. Additional TTE-level clinical context for the external test set is provided in **Supplemental Results 1.**

### 3.2. Validation of Morphologic Feature Detection

The proposed DL-based framework demonstrated robust performance in classifying the amount of pericardial effusion (normal, small, moderate, and large) (**Figure 2A**). In the internal test set, the model achieved per-class accuracies of 93.3% (normal), 81.8% (small), 85.8% (moderate), and 97.3% (large) (**Table 2**). F1-score of 0.933 (normal) and 0.864 (large) indicates particularly strong performance in detecting the extreme categories. In the external test set, performance remained consistent, with per-class accuracies of 94.2% (normal), small (83.6%), moderate (80.3%), and 92.3% (large) (**Table 2**). F1-score of 0.892 (normal) and 0.753 (large) reflects stable generalisation even across a separate institutional dataset. For pericardial effusion classification, the model also demonstrated excellent discriminative ability across severity thresholds (**Figure 3A**). In the internal test set, the AUROCs for detecting small or greater, moderate or greater, and large effusion were 0.95, 0.92, and 0.99, respectively. In the external test set, corresponding AUROCs were 0.98, 0.95, and 0.95, confirming stable performance across datasets.

**Figure 2.**
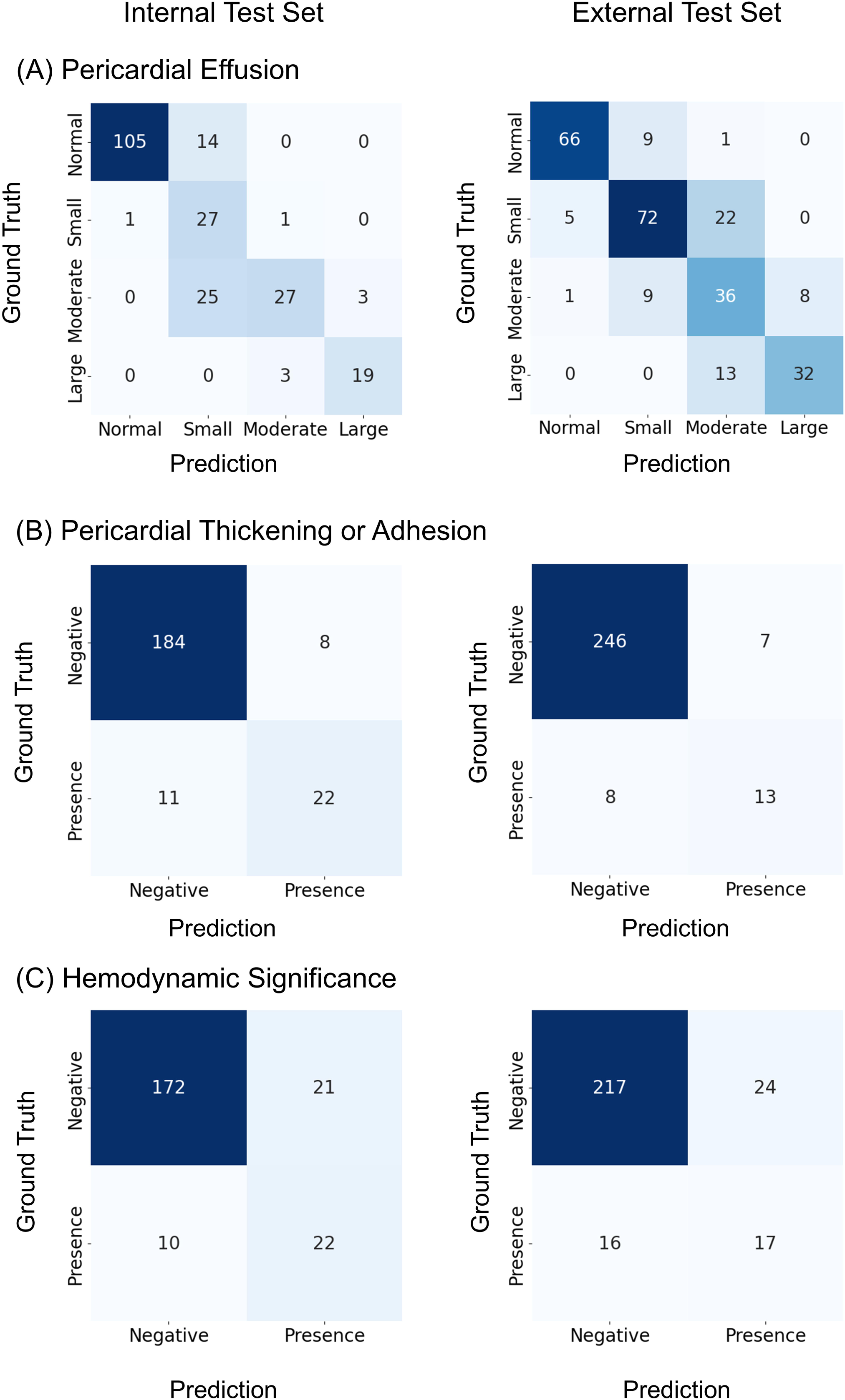
Confusion Matrices for Internal and External Test Sets Across Three Diagnostic Tasks: (A) Pericardial Effusion Classification, (B) Pericardial Thickening or Adhesion Detection, and (C) Hemodynamic Significance Assessment.

**Figure 3.**
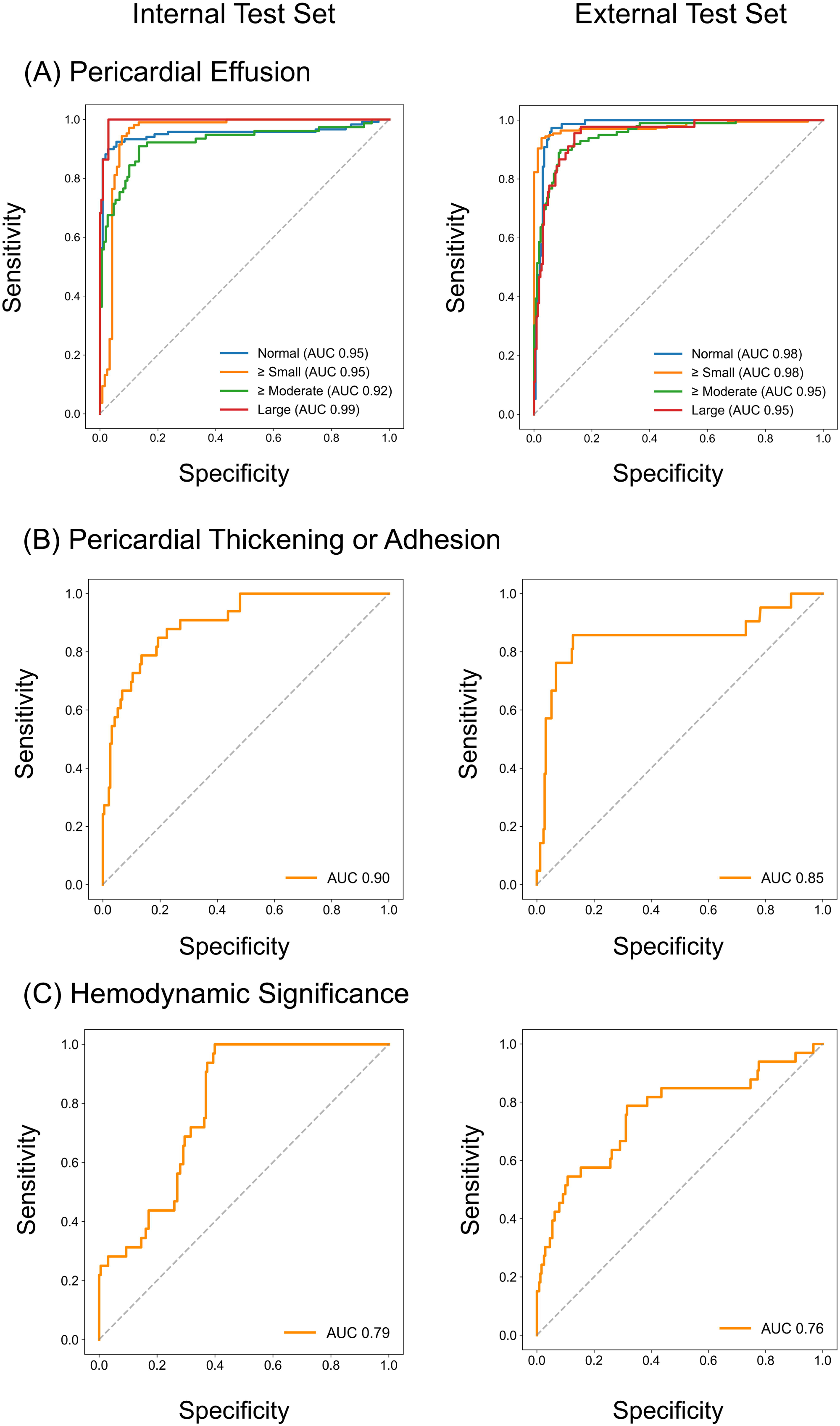
Receiver Operating Characteristics Curves for Internal and External Test Sets Across Three Diagnostic Tasks: (A) Pericardial Effusion Classification, (B) Pericardial Thickening or Adhesion Detection, and (C) Hemodynamic Significance Assessment.

**Table 2.** Diagnostic Performance of the Proposed DL-Based Framework Across Internal and External Test Datasets.

|  | Internal Test Dataset |  |  |  |  | External Test Dataset |  |  |  |  |
| --- | --- | --- | --- | --- | --- | --- | --- | --- | --- | --- |
|  | Accuracy | Precision | Sensitivity | Specificity | F1-score | Accuracy | Precision | Sensitivity | Specificity | F1-score |
| **Pericardial Effusion** |  |  |  |  |  |  |  |  |  |  |
| Normal | 0.933 | 0.991 | 0.882 | 0.991 | 0.933 | 0.942 | 0.917 | 0.868 | 0.970 | 0.892 |
| Small | 0.818 | 0.410 | 0.931 | 0.801 | 0.568 | 0.836 | 0.800 | 0.727 | 0.897 | 0.762 |
| Moderate | 0.858 | 0.871 | 0.491 | 0.977 | 0.628 | 0.803 | 0.500 | 0.667 | 0.836 | 0.571 |
| Large | 0.973 | 0.864 | 0.864 | 0.985 | 0.864 | 0.923 | 0.800 | 0.711 | 0.965 | 0.753 |
| **Pericardial Thickening/Adhesion** |  |  |  |  |  |  |  |  |  |  |
| Presence | 0.916 | 0.733 | 0.667 | 0.958 | 0.698 | 0.945 | 0.650 | 0.619 | 0.972 | 0.634 |
| **Hemodynamic Significance** |  |  |  |  |  |  |  |  |  |  |
| Presence | 0.862 | 0.512 | 0.688 | 0.891 | 0.587 | 0.855 | 0.415 | 0.515 | 0.904 | 0.460 |

Accuracy = correct classification rate for each diagnostic category; Precision = positive predictive value; Sensitivity = recall (true positive rate); Specificity = true negative rate; F1-score = harmonic mean of precision and sensitivity

In addition to pericardial effusion classification, the model’s performance in detecting pericardial thickening or adhesion was also evaluated. The optimal cutoff probability, determined from the internal validation set, was 0.107. Using this threshold, the model achieved an accuracy of 91.6% in the internal test set, with a precision of 73.3%, a sensitivity of 66.7%, a specificity of 95.8%, and an F1-score of 0.698 (**Table 2**, **Figure 2B**). Although sensitivity was moderate, the high specificity indicates that the model effectively minimises false positives and can provide reliable clinical support for detecting pericardial thickening and adhesion. In the external test set, the model achieved an accuracy of 94.5%, precision of 65.0%, sensitivity of 61.9%, specificity of 97.2%, and an F1-score of 0.634 (**Table 2**, **Figure 2B**). The AUROC for detecting pericardial thickening or adhesion was 0.90 in the internal test set and 0.85 in the external test set (**Figure 3B**), indicating good and consistent discriminative ability across datasets.

### 3.3. Validation of Hemodynamic Significance Detection

For the detection of hemodynamic significance, stepwise performance was evaluated according to the type of input provided (**Supplemental Results 2**). As additional hemodynamic information was sequentially incorporated—first Doppler measurements, then IVC measurements—diagnostic performance progressively improved. Specifically, the accuracy increased from 71.6% with B-mode videos only to 86.2% with the final input configuration including Doppler and IVC data in the internal test set. In parallel, the F1-score improved from 0.256 to 0.587. Notably, the model’s sensitivity increased from 33.4% to 68.8% as more functional information was provided. The AUROC also improved stepwise, from 0.70 with B-mode videos only, to 0.74 with the addition of Doppler measurements, and to 0.76 with further inclusion of IVC measurements in the internal test set. Similar trends were observed in the external test set. The accuracy increased from 76.3% to 85.5%, the F1-score improved from 0.253 to 0.460, and sensitivity increased from 33.3% to 51.5%. The AUROC also improved from 0.70 to 0.74 and finally to 0.76. The final model performance in the internal and external test sets is summarised in **Figure 2C**, **Figure 3C**, and **Table 2**.

### 3.4. Subgroup Analysis Based on Image Quality and Data Completeness

We evaluated the effect of IQ and data completeness on model performance within the internal test set. When stratified by IQ, performance declined in the poor-IQ group (**Supplemental Results 3**). Sensitivity for detecting pericardial thickening/adhesion and hemodynamic significance dropped to 45.5% and 16.7% respectively, compared to 80.0% and 77.8% in the good-IQ group. The AUROC for hemodynamic significance was also lower (0.68 vs. 0.81). After excluding the 42 poor-IQ cases, sensitivity improved from 66.7% to 77.3% for pericardial thickening/adhesion and from 68.8% to 80.8% for hemodynamic significance (**Table 3**). Confusion matrices and ROC curves for the good/fair IQ subgroup are shown **Figure 4**.

**Figure 4.**
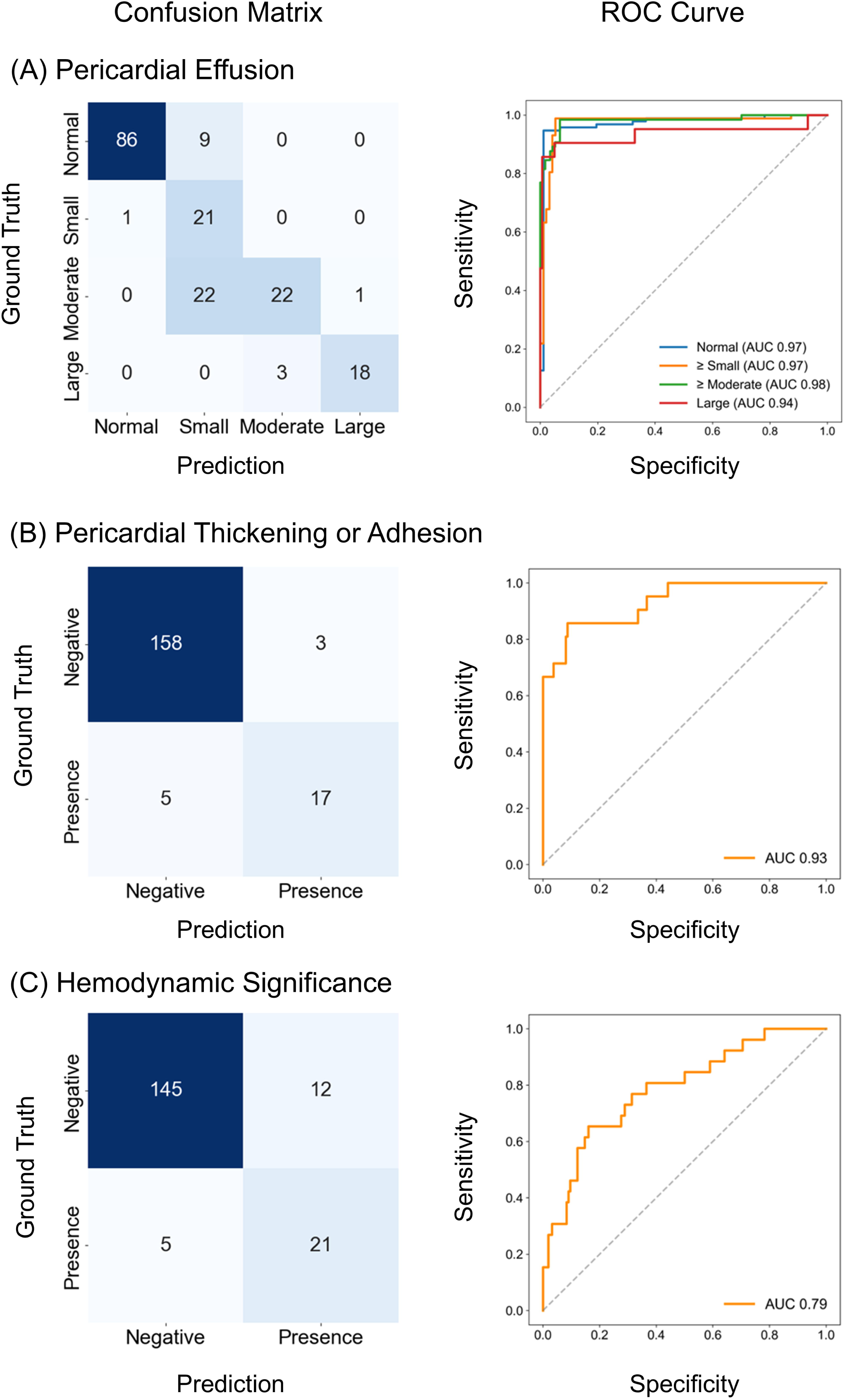
Diagnostic Performance of the DL-Based Framework in the Good/Fair IQ Subgroups of the Internal Test Set. Confusion matrices (left) and Receiver Operating Characteristics Curves (right) are presented for the three diagnostic tasks: A) Pericardial Effusion Classification, (B) Pericardial Thickening or Adhesion Detection, and (C) Hemodynamic Significance Assessment.

**Table 3.** Diagnostic Performance of the Proposed DL-Based Framework According to Image Quality in the Internal Test Set.

| IQ - Good, Fair (N = 183) |  |  |  |  |  |
| --- | --- | --- | --- | --- | --- |
|  | Accuracy | Precision | Sensitivity | Specificity | F1-score |
| **Pericardial Effusion** |  |  |  |  |  |
| Normal | 0.945 | 0.989 | 0.905 | 0.989 | 0.945 |
| Small | 0.825 | 0.404 | 0.955 | 0.807 | 0.568 |
| Moderate | 0.858 | 0.880 | 0.489 | 0.978 | 0.629 |
| Large | 0.978 | 0.947 | 0.857 | 0.994 | 0.900 |
| **Pericardial Thickening/Adhesion** |  |  |  |  |  |
| Presence | 0.956 | 0.850 | 0.773 | 0.981 | 0.810 |
| **Hemodynamic Significance** |  |  |  |  |  |
| Presence | 0.907 | 0.636 | 0.808 | 0.924 | 0.712 |

Data completeness showed a similar effect. When all five target B-mode views were available, sensitivity increased from 0.500 to 0.762 for detecting pericardial thickening/adhesion and from 0.571 to 0.778 for detecting hemodynamic significance (**Supplemental Results 4**). Performance also improved with a greater number of available B-mode clips: sensitivity increased from 0.500 to 0.704 for detecting pericardial thickening/adhesion and from 0.625 vs. 0.708 for hemodynamic significance when comparing cases with ≤7 available B-mode clips (lower tertile) to those with >7 clips (upper two tertiles) (**Supplemental Results 5)**.

### 3.5. Model Explainability

Grad-CAM^14^ visualisations were generated to interpret the model’s focus (**Figure 5**). In the first stage, the model for detecting pericardial effusion and pericardial thickening/adhesion predominantly focused on the pericardial region, suggesting the model appropriately localised features relevant to structural assessment. In contrast, in the second stage, the model for assessing hemodynamic significance demonstrated attention not only to the pericardium but also to intracardiac regions, indicating that the model simultaneously considered structural and functional information. This multi-regional focus was consistently across datasets.

**Figure 5.**
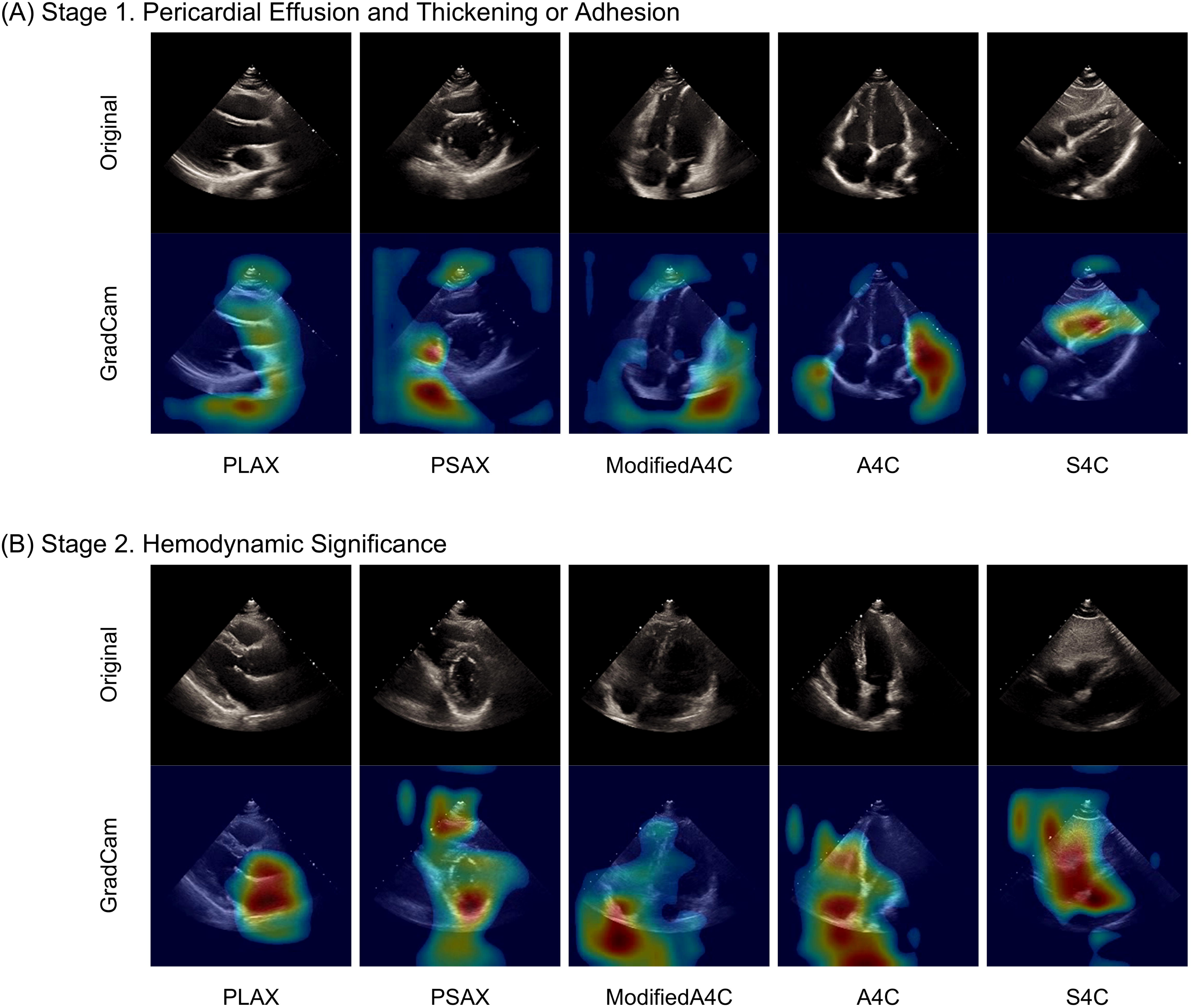
GRAD-CAM Visualization of Selected Cases from Internal and External Test Sets Across Two Diagnostic Stages: (A) Stage 1: Pericardial Effusion and Thickening or Adhesion, and (B) Stage 2: Hemodynamic Significance.

## 4. DISCUSSIONS

In this study, we developed and validated a DL-based framework for the automated identification of a broader spectrum of pericardial abnormalities using a limited set of standard TTE views. Unlike prior studies that have focused predominantly on detecting pericardial effusion^15–17^, our framework extends beyond effusion quantification to include evaluation of pericardial thickening, adhesion, and the hemodynamic significance associated with pericardial abnormalities. To achieve this, we adopted a two-stage approach that mirrors the clinical reasoning process, first assessing morphological features and then estimating the likelihood of hemodynamic impact. Furthermore, the proposed framework was validated not only on an internal test set but also on an independent external dataset from a distinct institution, demonstrating consistent performance.

Pericardial disease is common and presents with a broad spectrum of clinical phenotypes^1,2^. Pericardial effusion is the most frequently encountered manifestation, and it is thus unsurprising that most existing AI-based models have focused on detecting effusion and providing semi-quantitative estimation of its amount^15–17^. However, pericardial effusion represents only a portion of the overall disease continuum, and its clinical trajectory can vary significantly^1,2^. Some effusions may spontaneously resolve, whereas others rapidly accumulate and result in life-threatening tamponade. Even when tamponade is successfully managed, persistent inflammation may lead to constrictive pericarditis. Consequently, the absence of effusion cannot be equated with the absence of disease—a common source of misdiagnosis in clinical practice and a critical limitation of many existing AI-based approaches, which overlook more severe and clinically significant conditions. To our knowledge, this study represents the first attempt to develop an AI model capable of evaluating the full spectrum of pericardial disease, encompassing not only morphological features such as effusion, thickening, and adhesion, but also the functional consequences reflected in hemodynamic compromise. By implementing a two-stage architecture that mirrors clinical diagnostic reasoning, our framework introduces a novel paradigm that integrates structural and physiological assessment. This approach not only enables comprehensive disease characterisation but also supports informed clinical decision-making regarding the urgency and necessity of therapeutic intervention.

To enhance diagnostic robustness, the model incorporated a multi-view input strategy that leveraged complementary TTE views rather than relying on a single acoustic window. This design emulates real-world clinical practice, where multiple views are routinely obtained to ensure comprehensive evaluation of the pericardium. Furthermore, when multiple video clips were available for the same view, the model was designed to utilise all redundant inputs, reflecting the clinical tendency to acquire repeated images when needed for diagnostic confidence. Technically, these design principles were realised through (1) an explicit two-stage architecture that sequentially integrates morphological and functional features; (2) a multi-instance inference strategy that aggregates predictions across all feasible combinations of views and clips; and (3) uncertainty-weighted loss optimisation to account for aleatoric uncertainty and balance multi-task learning. Collectively, these strategies enhanced the model’s robustness and clinical applicability by emulating real-world diagnostic workflows while accommodating the inherent data variability in echocardiographic data.

Nevertheless, as with human interpretation, B-mode TTE videos alone were insufficient to achieve optimal performance for detecting hemodynamic significance. Incorporation of Doppler measurements and IVC features substantially improved diagnostic performance in a stepwise manner. However, several limitations remain. For instance, respiratory variation of the transmitral and transtricuspid inflow was not included in this study, and TDI was limited to septal motion without incorporating lateral annular movement for comparison. These limitations reflect the clinical reality that such advanced Doppler acquisitions were available only in a minority of cases, typically when tamponade or constrictive physiology was clinically suspected. Therefore, selectively incorporating these features when available may inadvertently introduce bias, as their presence itself signals operator suspicion. Still, in real-world AI deployment, leveraging such information when available could enhance both diagnostic accuracy and clinical utility. In addition, IVC features were derived exclusively from B-mode subcostal views, precluding analysis in cases with only M-mode acquisitions. To address this, we are developing an automated M-mode analysis module that will enable the broader inclusion of IVC assessments and improve compatibility with diverse acquisition protocols across institutions. This addition has the potential to improve not only model performance but also generalizability.

By integrating functional assessment with structural findings, our framework aims to provide a more comprehensive and clinically meaningful evaluation of pericardial disease. However, model performance for detecting hemodynamic significance remained relatively limited, particularly in terms of sensitivity and precision. While the restricted availability of Doppler and IVC data partially explains this, a more fundamental challenge lies in the intrinsic class imbalance in the developmental dataset. Despite incorporating multi-centre datasets, the number of patients exhibiting hemodynamic compromise remained small. We believe that expanding the training set with additional cases could help mitigate this imbalance and improve performance. At the same time, it is important to recognise that determining hemodynamic significance is a complex, physiology-based judgment that inherently involves a degree of subjectivity. Nevertheless, by ensuring expert-level labelling across all cases, we were able to train and validate a model capable of assessing not only morphological features but also the functional impact of pericardial disease, an advancement not previously demonstrated in prior AI-based studies.

While our framework demonstrated the capacity to assess both morphological and functional aspects of pericardial disease, its application in real-world clinical practice warrants caution. In this study, we deliberately included all test cases regardless of IQ or view completeness, allowing us to estimate the model’s expected performance under practical conditions. As shown in our subgroup analysis, diagnostic performance – particularly the sensitivity for detecting hemodynamic significance – declined in cases with poor IQ, missing target views, or limited numbers of available video clips. Importantly, these analyses were conducted as exploratory evaluations rather than predefined primary aims, and our in-house IQ labelling scheme, tailored to highlight pericardial structures, may limit generalizability to other datasets. Nevertheless, this issue is especially relevant in scenarios such as unstable or uncooperative patients, where comprehensive image acquisition may not be feasible. These findings highlight that, under suboptimal imaging conditions, the model’s performance may be compromised. However, this limitation is not unique to AI; even expert human interpretation would face similar challenges when presented with inadequate imaging data. In such situations, clinical decision-making must rely not solely on imaging but on the integration of patient history, symptoms, and physical examination. Thus, while the model offers valuable support in standard imaging scenarios, its outputs should be interpreted with clinical context in mind, particularly in technically challenging cases.

The present study has some limitations. First, although the developmental dataset was constructed from a multi-centre cohort and externally validated using data from an independent institution, all data originated from Korean institutions. Further validation using datasets from non-Korean populations is necessary to confirm generalizability across different ethnic and healthcare environments. Second, all datasets used in this study, including both developmental and external test sets, were retrospectively collected. Moreover, the study populations were not consecutively enrolled but were selectively sampled to collect a broad range of pericardial disease cases, supplemented with normal controls. As a result, the prevalence of pericardial disease was higher than that typically observed in routine echocardiographic practice. Nevertheless, such a retrospective design represents a critical and necessary step in the early development and validation of AI-based models for pericardial disease, a domain where automated analysis is still in its infancy. Future studies – either retrospective analyses of consecutively enrolled patients or prospective clinical trials - are warranted to evaluate the real-world clinical utility and integration of such models into routine diagnostic workflows. Finally, despite our efforts to minimise label noise by having all TTE examinations re-reviewed by experienced cardiologists, reference labelling based solely on TTE may be imperfect, given the inherent diagnostic challenges of pericardial disease. To address this, we are planning further validation using cases with confirmatory imaging, such as cardiac CT or MRI, to establish more definitive reference standards and assess model performance against these higher-fidelity benchmarks. Although the present framework was intentionally developed using a limited set of standard TTE views, we recognise that comprehensive evaluation of pericardial disease ultimately requires analysis of full echocardiographic examination. Several DL approaches are progressing toward this goal^22,23^. In parallel, we are developing an advanced model that incorporates the entire echocardiographic examination, supported by a substantially larger developmental dataset and a large consecutive cohort secured for external testing, which may help overcome the limitations of the current study and further strengthen AI-based pericardial assessment.

## Conclusions

We developed and externally validated a two-stage DL framework for the automated identification of major pericardial abnormalities using a limited set of standard TTE. Beyond effusion detection, the model provides broader morphologic assessment and offers supportive estimation of potential hemodynamic significance. While the model demonstrated consistent overall performance across internal and external test sets, sensitivity was affected by IQ and view completeness, underscoring the need for cautious interpretation in suboptimal imaging conditions. Within these constraints, the proposed framework may assist in flagging important pericardial findings and supporting timely referral for expert evaluation in routine clinical practice.

## Data Availability

The AI-Hub dataset used in this study may be available upon proper request and approval of a formal proposal. The external test dataset cannot be made publicly shared due to ethical restrictions imposed by the IRB of the study institution, as public access could compromise patient confidentiality and privacy. Researchers interested in accessing the minimal anonymized dataset may contact the corresponding author for further information.

## Contributors

**Sihyeon Jeong:** Conceptualisation, Methodology, Data curation, Formal Analysis

**In Tae Moon**: Validation, Investigation, Data Curation, Echocardiographic review and annotation

**Jaeik Jeon:** Software, Methodology, Resources

**Dawun Jeong:** Data Curation, Methodology

**Jina Lee**: Investigation, Project administration

**Jiyeon Kim**: Software, Model Training, Visualisation

**Seung-Ah Lee**: Investigation, Supervision, Echocardiographic review and annotation

**Yeonggul Jang:** Software supervision, Technological methodology oversight

**Yeonyee E. Yoon**: Conceptualisation, Supervision, Echocardiographic review and annotation, Clinical interpretation, Writing, Review and Editing

**Hyuk-Jae Chang**: Conceptualisation, Supervision, Funding acquisition All authors read and approved the final version of the manuscript.

## Acknowledgments

This work was supported by a grant from the Institute of Information & Communications Technology Planning & Evaluation (IITP) funded by the Korea government (Ministry of Science and ICT) (No.2022000972, Development of a Flexible Mobile Healthcare Software Platform Using 5G MEC); and the Medical AI Clinic Program through the NIPA funded by the MSIT. (Grant No.: H0904-24-1002).

## Declaration of Interests

J.J., J.K., S.A.L., Y.J., and Y.E.Y. are currently affiliated with Ontact Health, Inc. Y.E.Y., and H.J.C. hold stock in Ontact Health, Inc. The other authors have no conflicts of interest to declare.

## Data Sharing Statement

The AI-Hub dataset used in this study may be available upon proper request and approval of a formal proposal. The external test dataset cannot be made publicly shared due to ethical restrictions imposed by the IRB of the study institution, as public access could compromise patient confidentiality and privacy. Researchers interested in accessing the minimal anonymised dataset may contact the corresponding author for further information.

## Supplemental Methods

**Supplemental Methods 1. Study Population**

**Supplemental Methods 2. Diagnostic Classification Logic for Pericardial Disease Category**

**Supplemental Methods 3. Proposed Image Quality (IQ) Criteria for Deep Learning-Based Pericardial Disease Assessment**

**Supplemental Methods 4. Two-Stage Deep Learning Framework Details**

**Supplemental Methods 5. Comparison of Inference Strategies for Multi-View Redundancy Handling**

**Supplemental Methods 6. IVC Segmentation Module for Diameter and Respiratory Variation Estimation**

## Supplemental Results

**Supplemental Results 1. External Test Set Characteristics (Per-TTE)**

**Supplemental Results 2. Stepwise Performance of the Hemodynamic Significance Detection Model According to Input Configuration**

**Supplemental Results 3. Performance by Image Quality Subgroup**

**Supplemental Results 4. Model Performance Stratified by Availability of All Five Target B-Mode Views**

**Supplemental Results . Model Performance Stratified by Number of Available B-mode Video Clips (Upper Two Tertiles vs. Lower Tertile)**

**References 18-21**

---

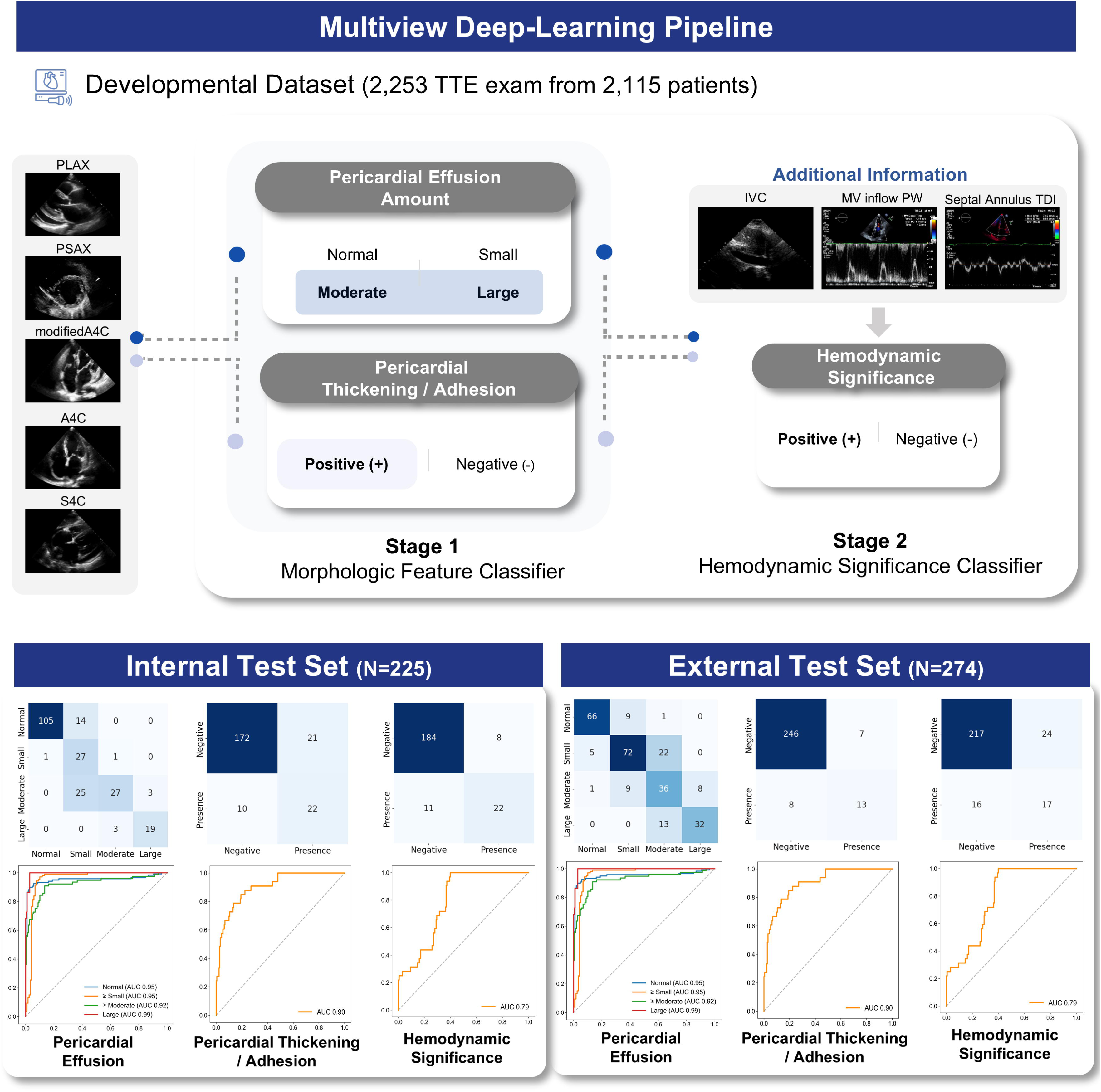

## Notes

### Competing Interest Statement

J.J., J.K., S.A.L., Y.J., and Y.E.Y. are currently affiliated with Ontact Health, Inc. Y.E.Y., and H.J.C. holds stock in Ontact Health, Inc. The other authors have no conflicts of interest to declare.

### Author Declarations

Institutional Review Board of Yonsei University Health System, Seoul National University Hospital, Chungnam National University Hospital, Hanyang University Hospital, and Soonchunhyang University Bucheon Hospital gave ethical approval for this work and waived the requirement for informed consent (protocol Nos. 2021-0147-003, CNUH 2021-04-032, HYUH 2021-03-026-003, SCHBC 2021-03-007-001, B 2104/677-004). In addition, Institutional Review Board of Uijeongbu Eulji University Hospital gave ethical approval for the external test dataset (protocol No. 2025-03-018).

### Summary of Updates

This version includes clarifications of the study scope and revisions for improved clarity and readability across the manuscript. No changes were made to the scientific content or conclusions.

