## Supplemental material for "Deep Learning-Based Multi-View Echocardiographic Framework for Comprehensive Diagnosis of Pericardial Disease"

**Supplemental Methods 1. Study Population**

The AI-Hub dataset^4^ comprises 30,000 transthoracic echocardiographic (TTE) examinations retrospectively collected from five tertiary hospitals in South Korea, including Chungnam National University Hospital, Hanyang University Hospital, Seoul National University Bundang Hospital, Severance Hospital, and Soonchunhyang University Seoul Hospital, between 2012 and 2021. The dataset includes a broad spectrum of cardiovascular disease categories, ranging from normal findings to ischemic heart disease, cardiomyopathy, pulmonary hypertension and embolism, pericardial disease, valvular heart disease, cardiac mass, and congenital heart disease.

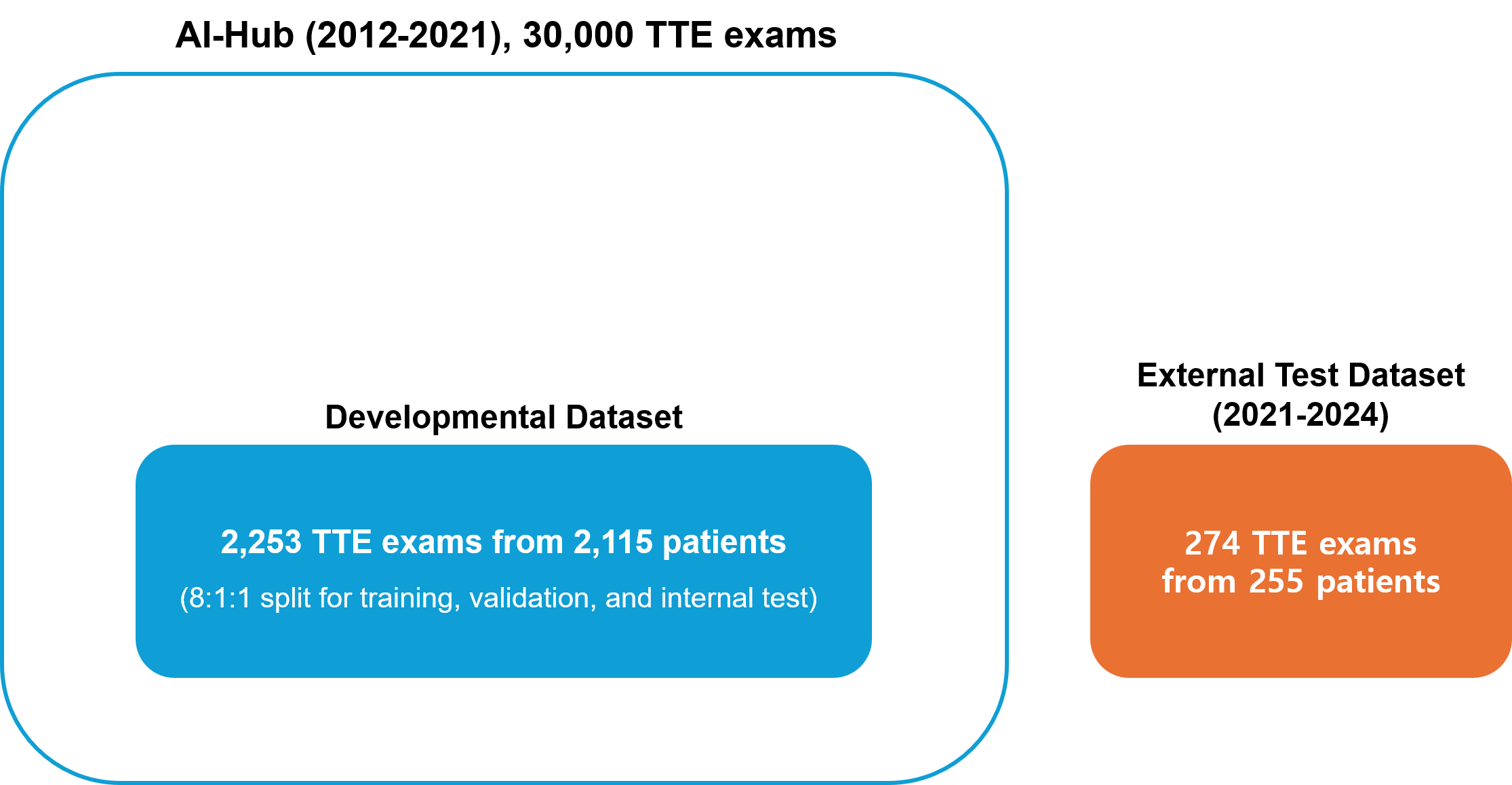

*For the present study, cases of normal pericardium were drawn from the “normal” category. These subjects were asymptomatic individuals undergoing health check-up TTE examinations, and only those with normal pericardial findings were included. Pericardial disease cases were extracted from the “pericardial disease” category, identified using ICD-10 diagnostic codes corresponding to pericardial conditions (I30.9: acute pericarditis, unspecified; I31.1: chronic constrictive pericarditis; I31.3: pericardial effusion, noninflammatory; I31.9: disease of pericardium, unspecified; C79.88: secondary malignant neoplasm with pericardial involvement). Only cases with matching echocardiographic reports confirming the diagnosis of pericardial disease were retained to reduce potential data contamination.*

**Supplemental Methods 2. Diagnostic Classification Logic for Pericardial Disease Category**

| **Pericardial Effusion** | **Pericardial Thickening/Adhesion** | **Hemodynamic Significance** | **Disease Category** |
| --- | --- | --- | --- |
| - | - | - | Normal Pericardium |
| + | - | - | Pericardial Effusion |
| + | - | + | Tamponade |
| + | + | - | Pericarditis with Effusion, but without Hemodynamic Significance |
| + | + | + | Effusive Constrictive Pericarditis |
| - | + | - | Pericarditis without Hemodynamic Significance |
| - | + | + | Constrictive Pericarditis |

**Supplemental Methods 3. Proposed Image Quality (IQ) Criteria for Deep Learning-Based Pericardial Disease Assessment**

| Grade | Definition | Key Criteria |
| --- | --- | --- |
| **Good** | The pericardium is clearly visualized throughout the cardiac cycle, and key anatomical structures required for the assessment of effusion, adhesion, or thickening are well delineated. | - Sharp visualization of the pericardium and adjacent epicardial fat - Clear separation between the pericardium and LV/RV free walls - Pericardial effusion (if present) is continuously visible - Minimal artifacts in Doppler and IVC views |
| **Fair** | Overall evaluable, but with partial limitations due to suboptimal visualization or artifacts affecting the interpretation of pericardial structures. | - Partial discontinuity or dropout of pericardial borders - Difficulty in confidently identifying loculated effusion - Mild artifacts or foreshortening affecting Doppler or IVC analysis |
| **Poor** | The pericardium and related structures are poorly visualized, making the evaluation of effusion, adhesion, or hemodynamic significance unreliable. | - Diffuse non-visualization or obscuration of the pericardium - Significant shadowing, rib artifact, or low-gain settings - IVC view not segmentable due to image degradation - Cardiac motion poorly captured (e.g., severe foreshortening or image dropout) |

**Supplemental Methods 4. Two-Stage Deep Learning Framework Details**

**4.1. Morphological Feature Modeling**

Each TTE views – parasternal long-axis (PLAX), parasternal short-axis (PSAX), apical four-chamber (A4C), A4C right ventricular (RV) focused or modified, subcostal four-chamber (SC4C) - was processed independently using 16–32 frame clips extracted from a full cardiac cycle. Cardiac cycles were segmented using electrocardiogram (ECG) timing embedded in the DICOM files or manually verified frame markers when ECG metadata were unavailable. All frames were resized to 224×224 pixels and normalized to [-1, 1].

A modified R(2+1)D-18 backbone was used to extract spatiotemporal features from each view. To accommodate the variable numbers of available views per patient, extracted features were pooled using AdaptiveAvgPool3d and zero-padded prior to concatenation. The classification head was branched into two outputs: (1) a four-class softmax for pericardial effusion severity (normal, small, moderate, large) and (2) a binary classifier for pericardial thickening and/or adhesion. Task optimization followed the homoscedastic uncertainty-weighting scheme^16^. The total loss is

$$\mathcal{L=}e^{-s_{1}}\mathcal{L}_{effusion}+ e^{-s_{2}}\mathcal{L}_{thk/adh}+ s_{1}+ s_{2}$$

where $\mathcal{L}_{effusion}$ is label-smoothed cross-entropy^19^, $\mathcal{L}_{thk/adh}$ is focal loss^20^, and $s_{1}$, $s_{2}$ are learnable log-variance terms that automatically balance the two tasks according to their predictive uncertainty.

**4.2. Functional Feature Integration**

Functional inputs included (a) two binary flags derived from the IVC module (dilatation and plethora) and (b) six Doppler features extracted from mitral inflow pulsed-wave (PW) Doppler (E, A, deceleration time) and septal tissue Doppler imaging (TDI) (a’, e’, s’). These functional features were concatenated with the pooled embeddings from Stage 1 and passed to a final binary classifier to predict hemodynamic significance.

**4.3 Training Strategy**

Training was performed using the AdamW^21^ optimizer with an initial learning rate = 0.002, a batch size of 8 per GPU (effective batch size = 24 across 3 GPUs), for a total of 300 epochs. Loss functions included label-smoothed cross-entropy for multi-class effusion classification and focal loss for the binary classification of pericardial thickening/adhesion and hemodynamic significance. To enhance robustness, we applied echocardiography-specific data augmentation techniques, including spatial transformation (translation, rotation up to ±30°, scaling, flipping, shadowing) and temporal transformation (frame sequence reversal).

**Supplemental Methods 5. Comparison of Inference Strategies for Multi-View Redundancy Handling**

We compared three different inference strategies to determine the optimal approach for handling view redundancy. This comparison was performed using the internal validation set.

- **Method 1:** The best-quality sample for each view is selected based on the view quality score from the view classification process, and predictions are performed using the combination of these selected samples across views.
- **Method 2:** For each echocardiographic view, spatiotemporally aggregated features were first extracted from all available video clips. The features corresponding to the same view were then averaged at the feature level to produce a single representative feature vector per view. Finally, the representative features from each view were integrated to generate the final prediction.
- **Method 3:** Exhaustive combination-based probability aggregation, in which all possible multi-view combinations were evaluated to generate predictions, and final classification was determined by aggregating the predicted probabilities across all combinations.

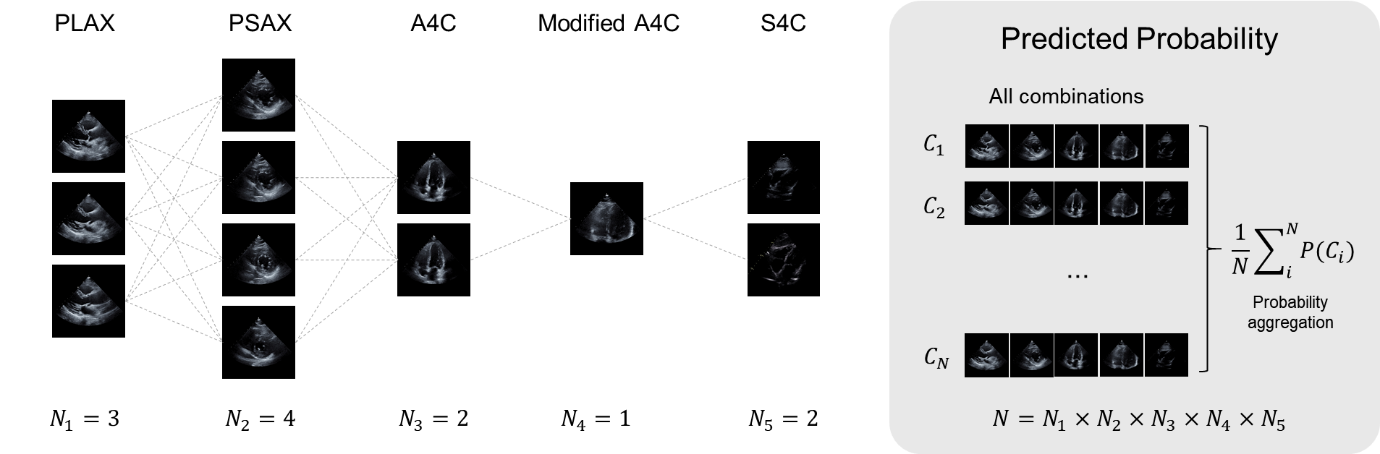

The example illustrates Method 3, in which multiple video clips were available for each echocardiographic view: PLAX (3 clips), PSAX (4 clips), A4C (2 clips), modified A4C (1 clip), and SC4C (2 clips). In this case, the total number of possible multi-view combinations was 3 × 4 × 2 × 1 × 2 = 48. Predictions were generated for each of the 48 combinations, and the final classification was determined by aggregating the predicted probabilities across all combinations.

**5.1. Confusion Matrices for Each Diagnostic Task Using Three Inference Strategies in the Internal Validation Set**

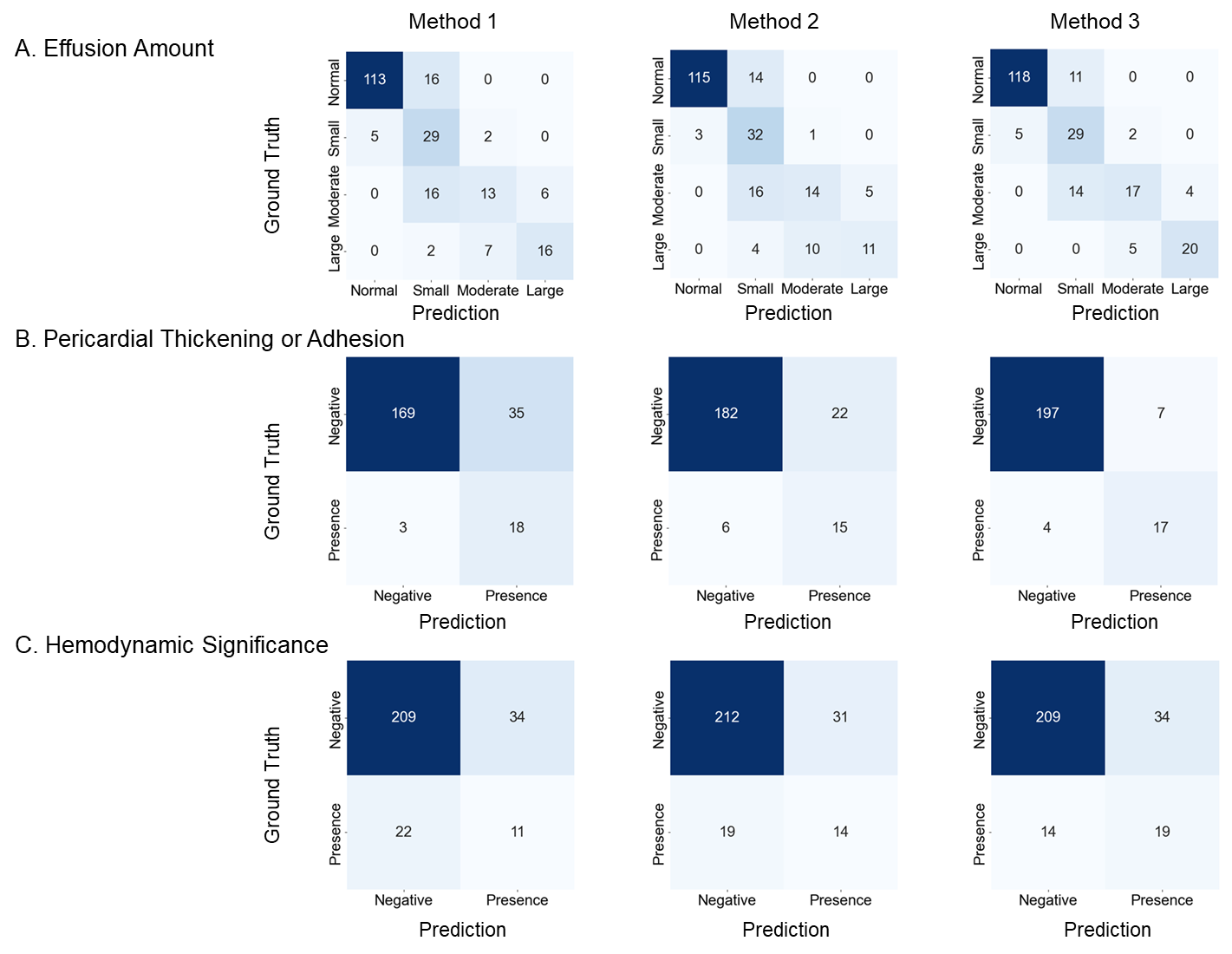

**5.2. Comparison of Inference Strategies for Handling Multiple Video Clips per View across Three Diagnostic Tasks in the Internal Validation Set**

|  | **Method 1** | | | | | | **Method 2** | | | | | | | | | | **Method 3** | | | | | | | |
| --- | --- | --- | --- | --- | --- | --- | --- | --- | --- | --- | --- | --- | --- | --- | --- | --- | --- | --- | --- | --- | --- | --- | --- | --- |
|  | **Acc** | **Prec** | **Sen** | **Spe** | **F1-score** | | **Acc** | | **Prec** | | **Sen** | | **Spe** | | **F1-score** | | **Acc** | | **Prec** | | **Sen** | | **Spe** | **F1-score** |
| **Pericardial Effusion** | | | | | | | | | | | | | | | | | | | | | | | | |
| Normal | 0.907 | 0.958 | 0.877 | 0.948 | 0.915 | | 0.924 | | 0.975 | | 0.892 | | 0.969 | | 0.931 | | 0.929 | | 0.960 | | 0.915 | | 0.948 | 0.937 |
| Small | 0.818 | 0.460 | 0.806 | 0.820 | 0.586 | | 0.831 | | 0.485 | | 0.889 | | 0.820 | | 0.628 | | 0.858 | | 0.537 | | 0.806 | | 0.868 | 0.644 |
| Moderate | 0.862 | 0.591 | 0.317 | 0.953 | 0.456 | | 0.858 | | 0.560 | | 0.400 | | 0.942 | | 0.467 | | 0.889 | | 0.708 | | 0.486 | | 0.963 | 0.576 |
| Large | 0.933 | 0.727 | 0.640 | 0.970 | 0.681 | | 0.916 | | 0.688 | | 0.440 | | 0.975 | | 0.537 | | 0.960 | | 0.833 | | 0.800 | | 0.980 | 0.816 |
| **Pericardial Thickening/Adhesion** | | | | | |  | |  | |  | |  | |  | |  | |  | |  | |  | |  |
| Presence | 0.876 | 0.405 | 0.714 | 0.892 | 0.517 | | 0.831 | | 0.340 | | 0.857 | | 0.828 | | 0.487 | | 0.951 | | 0.708 | | 0.810 | | 0.966 | 0.756 |
| **Hemodynamic Significance** | | | | | |  | |  | |  | |  | |  | |  | |  | |  | |  | |  |
| Presence | 0.819 | 0.311 | 0.424 | 0.872 | 0.360 | | 0.797 | | 0.244 | | 0.333 | | 0.860 | | 0.262 | | 0.826 | | 0.359 | | 0.576 | | 0.860 | 0.442 |

Abbreviations: Acc = Accuracy; Prec = Precision; Sen = Sensitivity; Spe = Specificity.

Performance metrics (accuracy, precision, sensitivity, specificity, and F1-score) are reported for each task and method. Method 3, based on exhaustive probability aggregations across all view combinations, demonstrated the most consistent performance.

**5.3. ROC Curves for Each Diagnostic Task Using Three Inference Strategies in the Internal Validation Set**

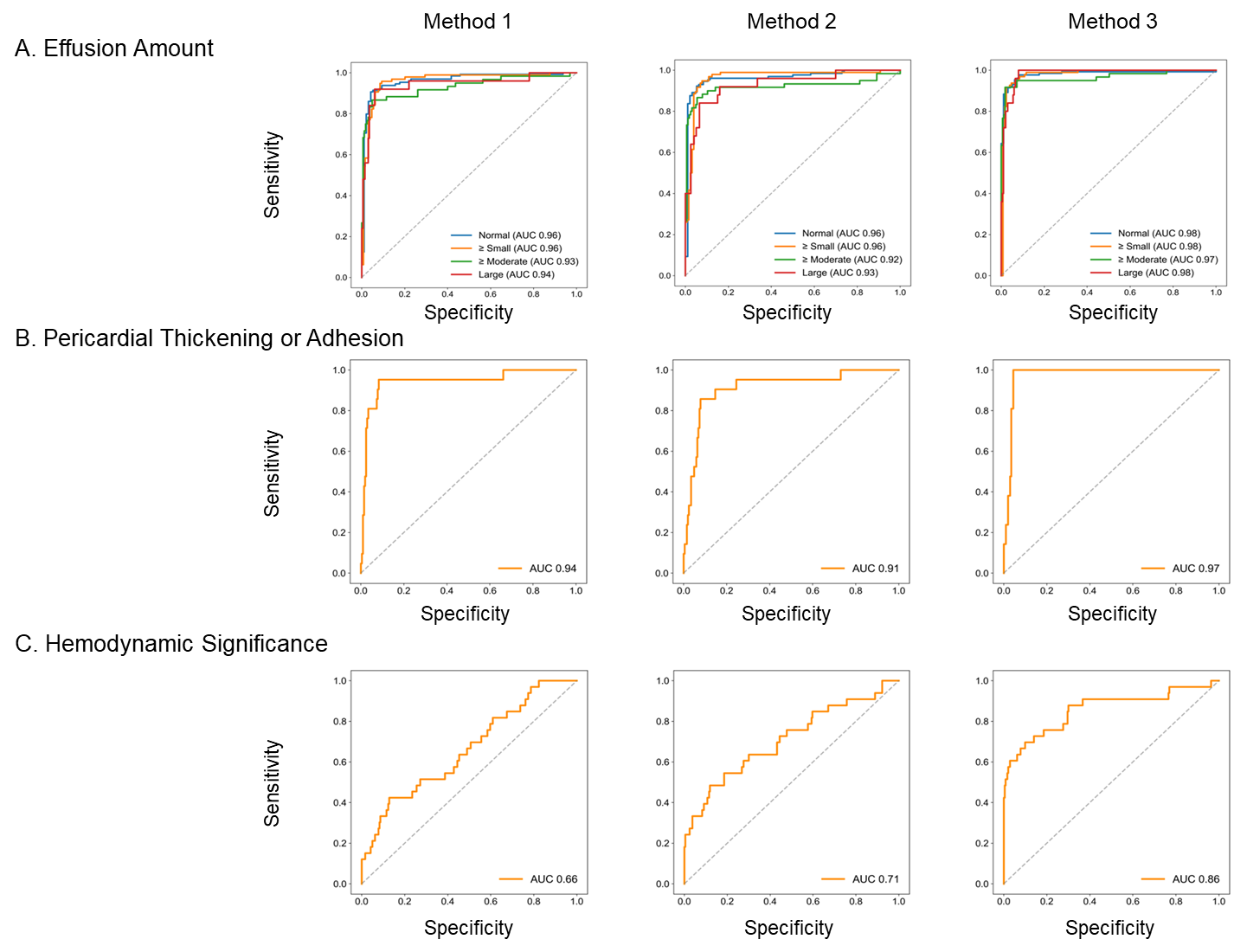

**Supplemental Methods 6. IVC Segmentation Module for Diameter and Respiratory Variation Estimation**

We have newly developed an artificial intelligence (AI)-based inferior vena cava (IVC) segmentation module to automatically estimate maximal IVC diameter and respiratory variation from full-length subcostal long-axis IVC cine loops. The model was built on a U-Net architecture with temporal context encoding and generates per-frame binary masks of the IVC lumen. The IVC diameter was calculated as the vertical axis length of the predicted segmentation mask. To improve measurement stability, a heatmap-guided regression head was integrated to smooth the diameter estimation, particularly in low-contrast frames. Respiratory variation was quantified using the collapsibility index $(D_{max}-D_{min})/D_{max}$ over the respiratory cycle.

Internal validation on an independent in-house dataset (n=51) demonstrated robust performance, with a mean Dice coefficient of 0.86 ± 0.03 for segmentation, a mean absolute error of 1.8 mm for diameter estimation, and classification accuracy of 0.91 and 0.89 for IVC dilatation and plethora detection, respectively.

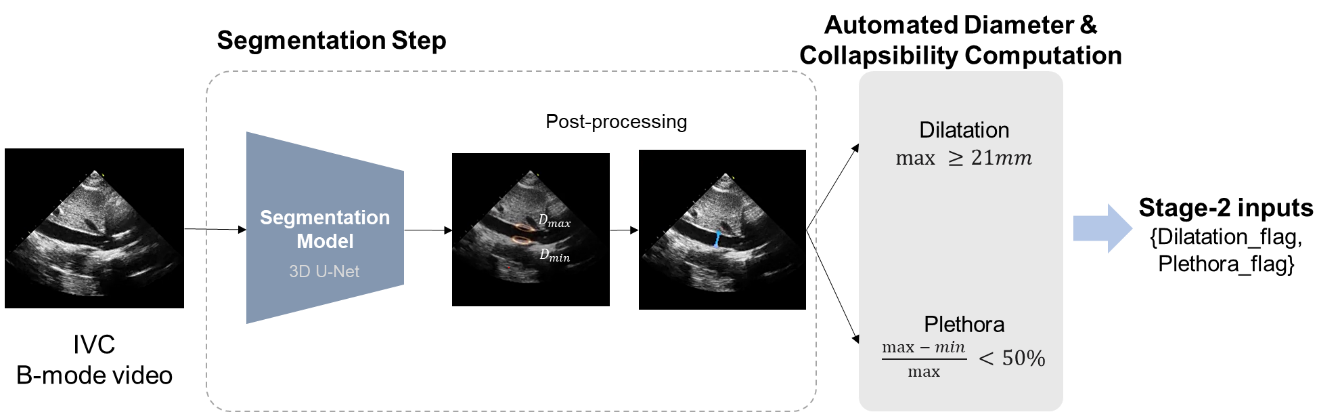

Pre-processing module that converts a subcostal IVC cine loop into two categorical cues for the downstream classifier. The 3D U-Net first delineates the IVC lumen on a frame-by-frame basis. Post-processing then extracts the largest and smallest diameters over the respiratory cycle and, from these, derives binary “dilatation” and “plethora” flags. Only these two indicators are forwarded to the multi-view fusion network.

**Supplemental Results 1. External Test Set Characteristics**

|  | **Normal** | **Pericardial disease** |
| --- | --- | --- |
|  | **(N = 65)** | **(N = 209)** |
| **Care setting at the time of TTE** |  |  |
| Outpatient | 65 (100%) | 43 (20.6%) |
| Inpatient | - | 154 (73.7%) |
| Emergency department | - | 12 (5.7%) |
| **Cardiac surgery related** |  |  |
| No | **-** | 201 (96.2%) |
| Yes | **-** | 8 (3.8%) |
| **Etiology** |  |  |
| Malignant | **-** | 22 (10.5%) |
| Tuberculous | **-** | 18 (8.6%) |
| Hemopericardium | **-** | 5 (2.4%) |
| Transudate | **-** | 30 (14.4%) |
| Idiopathic or Other | **-** | 134 (64.1%) |

**Supplemental Results 2. Stepwise Performance of the Hemodynamic Significance Detection Model According to Input Configuration**

Model performance was first evaluated using B-mode echocardiographic videos only, followed by the addition of Doppler measurements (mitral inflow PW Doppler and septal TDI), and finally by incorporating IVC measurements.

**2.1. Stepwise Confusion Matrices for Hemodynamic Significance Detection in Internal and External Test Sets**

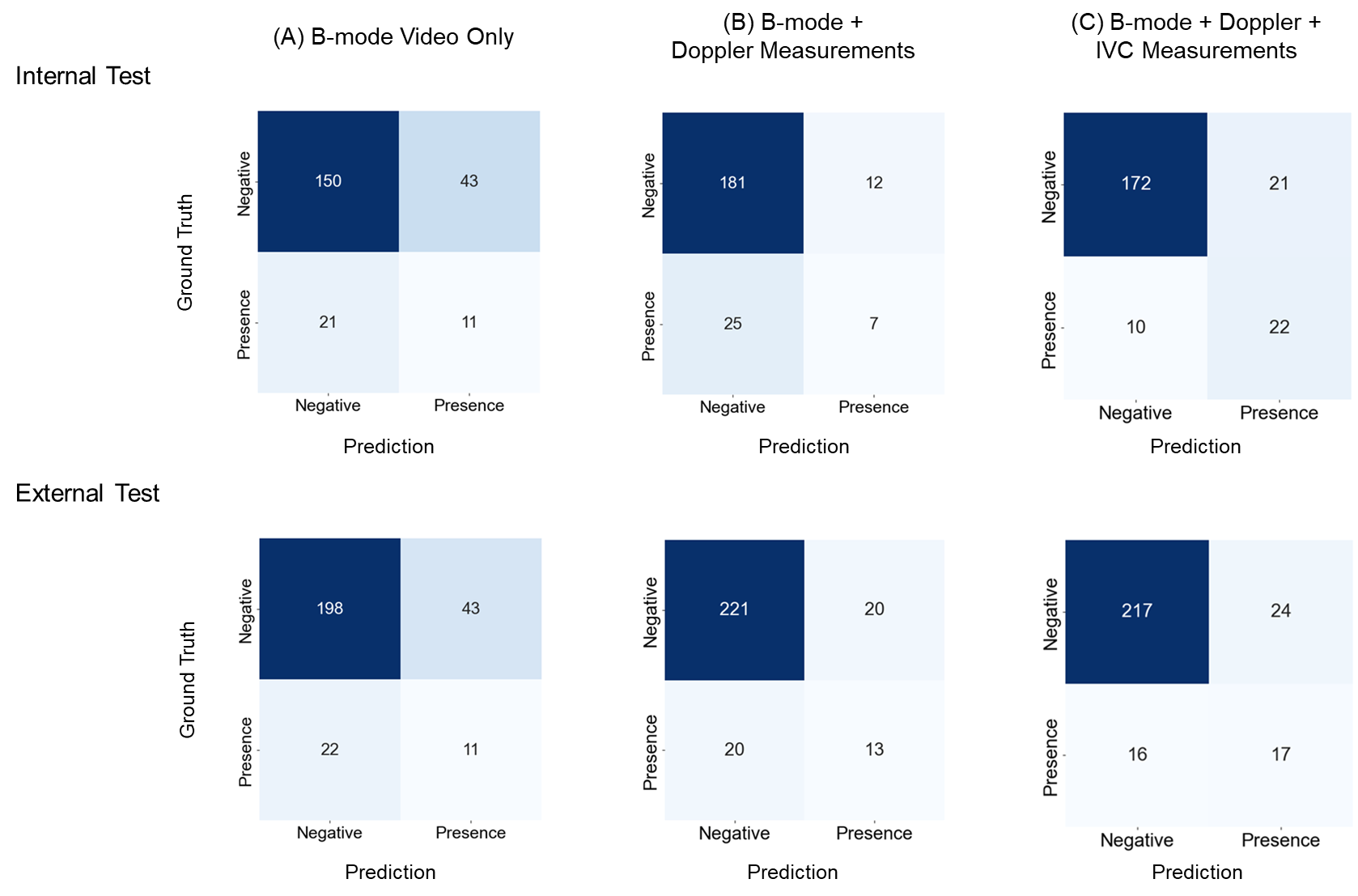

**2.2. Stepwise Diagnostic Performance Metrics for Hemodynamic Significance Detection In Internal and External Test Sets**

|  | **Internal Test Dataset** | | | | |  | **External Test Dataset** | | | | |
| --- | --- | --- | --- | --- | --- | --- | --- | --- | --- | --- | --- |
|  | **Accuracy** | **Precision** | **Sensitivity** | **Specificity** | **F1-score** |  | **Accuracy** | **Precision** | **Sensitivity** | **Specificity** | **F1-score** |
| **B-mode Only** | | | | | | | | | | | |
| Presence | 0.716 | 0.204 | 0.334 | 0.777 | 0.256 |  | 0.763 | 0.204 | 0.333 | 0.822 | 0.253 |
| **B-mode ＋ Doppler Measurements** | | | | | | | | | | | |
| Presence | 0.836 | 0.368 | 0.219 | 0.938 | 0.275 |  | 0.854 | 0.394 | 0.394 | 0.917 | 0.394 |
| **B-mode ＋ Doppler ＋ IVC Measurements** | | | | | | | | | | | |
| Presence | 0.862 | 0.512 | 0.688 | 0.891 | 0.587 |  | 0.855 | 0.415 | 0.515 | 0.904 | 0.460 |

**2.3. Stepwise Changes in AUROC for Hemodynamic Significance Detection Across Input Configuration**

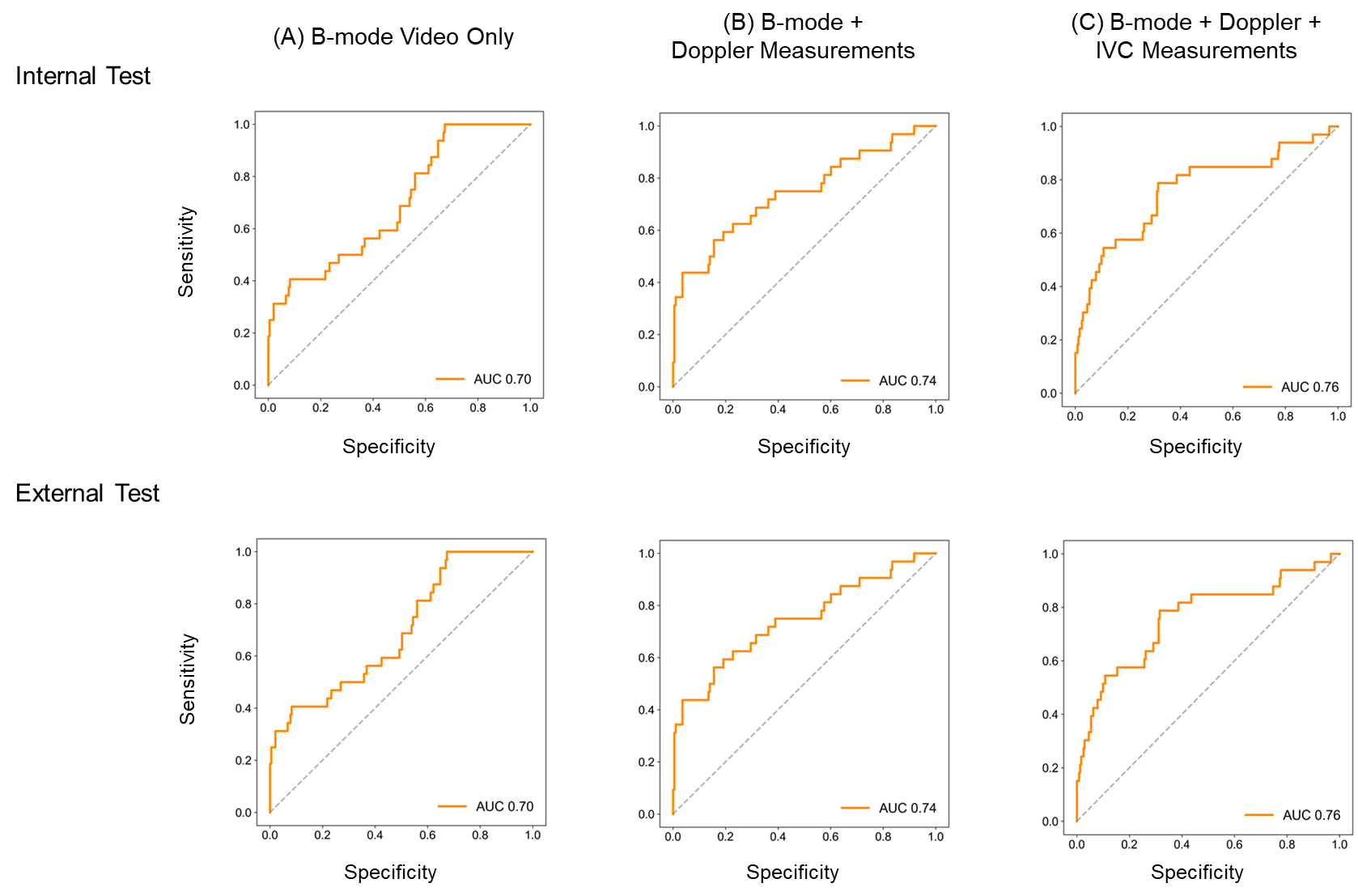

**Supplemental Results 3. Performance by Image Quality Subgroup**

**3.1. Confusion Matrices for Good, Fair, and Poor IQ Groups**

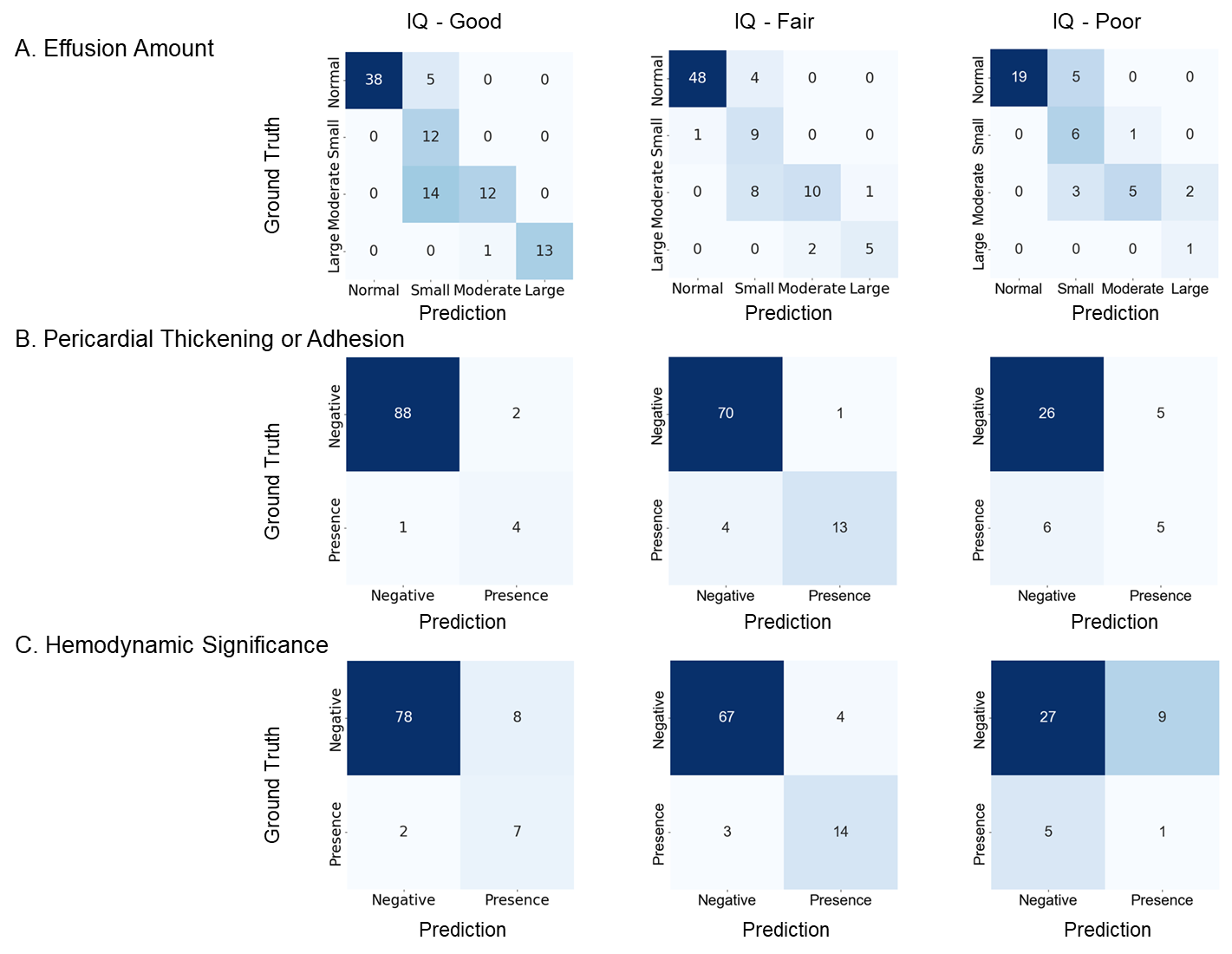

**3.2. Diagnostic Performance Metrics According to IQ**

|  | **IQ - Good (N = 95)** | | | | | | **IQ - Fair (N = 88)** | | | | | | | | | | **IQ - Poor (N = 42)** | | | | | | | |
| --- | --- | --- | --- | --- | --- | --- | --- | --- | --- | --- | --- | --- | --- | --- | --- | --- | --- | --- | --- | --- | --- | --- | --- | --- |
|  | **Acc** | **Prec** | **Sen** | **Spe** | **F1-score** | | **Acc** | | **Prec** | | **Sen** | | **Spe** | | **F1-score** | | **Acc** | | **Prec** | | **Sen** | | **Spe** | **F1-score** |
| **Pericardial Effusion** | | | | | | | | | | | | | | | | | | | | | | | | |
| Normal | 0.947 | 1.000 | 0.884 | 1.000 | 0.938 | | 0.943 | | 0.980 | | 0.923 | | 0.972 | | 0.951 | | 0.881 | | 1.000 | | 0.792 | | 1.000 | 0.884 |
| Small | 0.800 | 0.387 | 1.000 | 0.771 | 0.558 | | 0.852 | | 0.429 | | 0.900 | | 0.846 | | 0.581 | | 0.786 | | 0.429 | | 0.857 | | 0.771 | 0.571 |
| Moderate | 0.842 | 0.923 | 0.462 | 0.986 | 0.615 | | 0.875 | | 0.833 | | 0.526 | | 0.971 | | 0.645 | | 0.857 | | 0.833 | | 0.500 | | 0.969 | 0.625 |
| Large | 0.990 | 1.000 | 0.929 | 1.000 | 0.963 | | 0.966 | | 0.833 | | 0.714 | | 0.988 | | 0.769 | | 0.952 | | 0.333 | | 1.000 | | 0.951 | 0.500 |
| **Pericardial Thickening/Adhesion** | | | | | |  | |  | |  | |  | |  | |  | |  | |  | |  | |  |
| Presence | 0.968 | 0.667 | 0.800 | 0.978 | 0.727 | | 0.943 | | 0.929 | | 0.765 | | 0.986 | | 0.839 | | 0.738 | | 0.500 | | 0.455 | | 0.839 | 0.476 |
| **Hemodynamic Significance** | | | | | |  | |  | |  | |  | |  | |  | |  | |  | |  | |  |
| Presence | 0.895 | 0.467 | 0.778 | 0.907 | 0.583 | | 0.920 | | 0.778 | | 0.824 | | 0.944 | | 0.800 | | 0.667 | | 1.000 | | 0.167 | | 0.750 | 0.125 |

Abbreviations: Acc = Accuracy; Prec = Precision; Sen = Sensitivity; Spe = Specificity.

**3.3. ROC Curve Analysis Across IQ Subgroups**

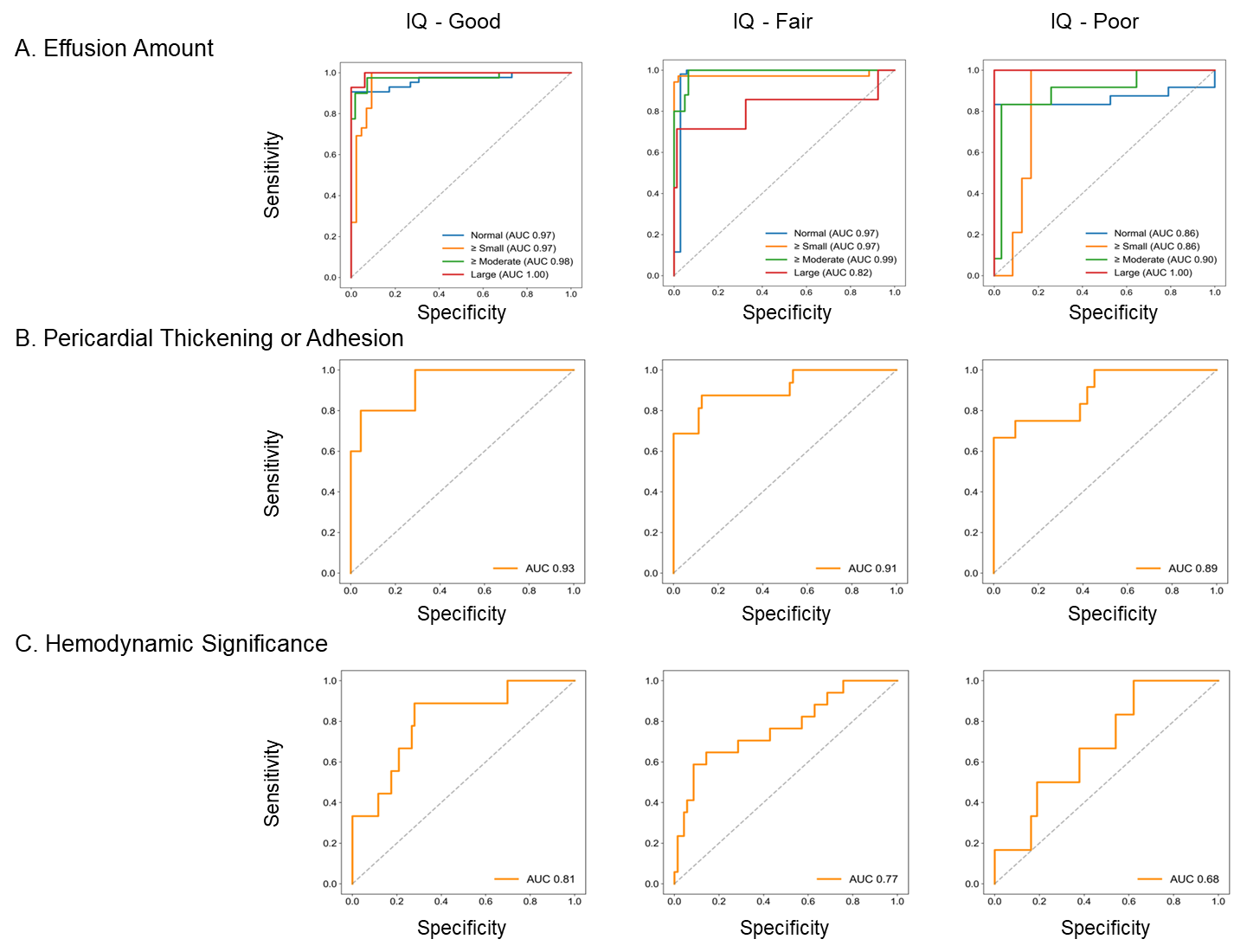

**Supplemental Results 4. Model Performance Stratified by Availability of All Five Target B-Mode Views**

**4.1. Confusion Matrices by View Completeness**

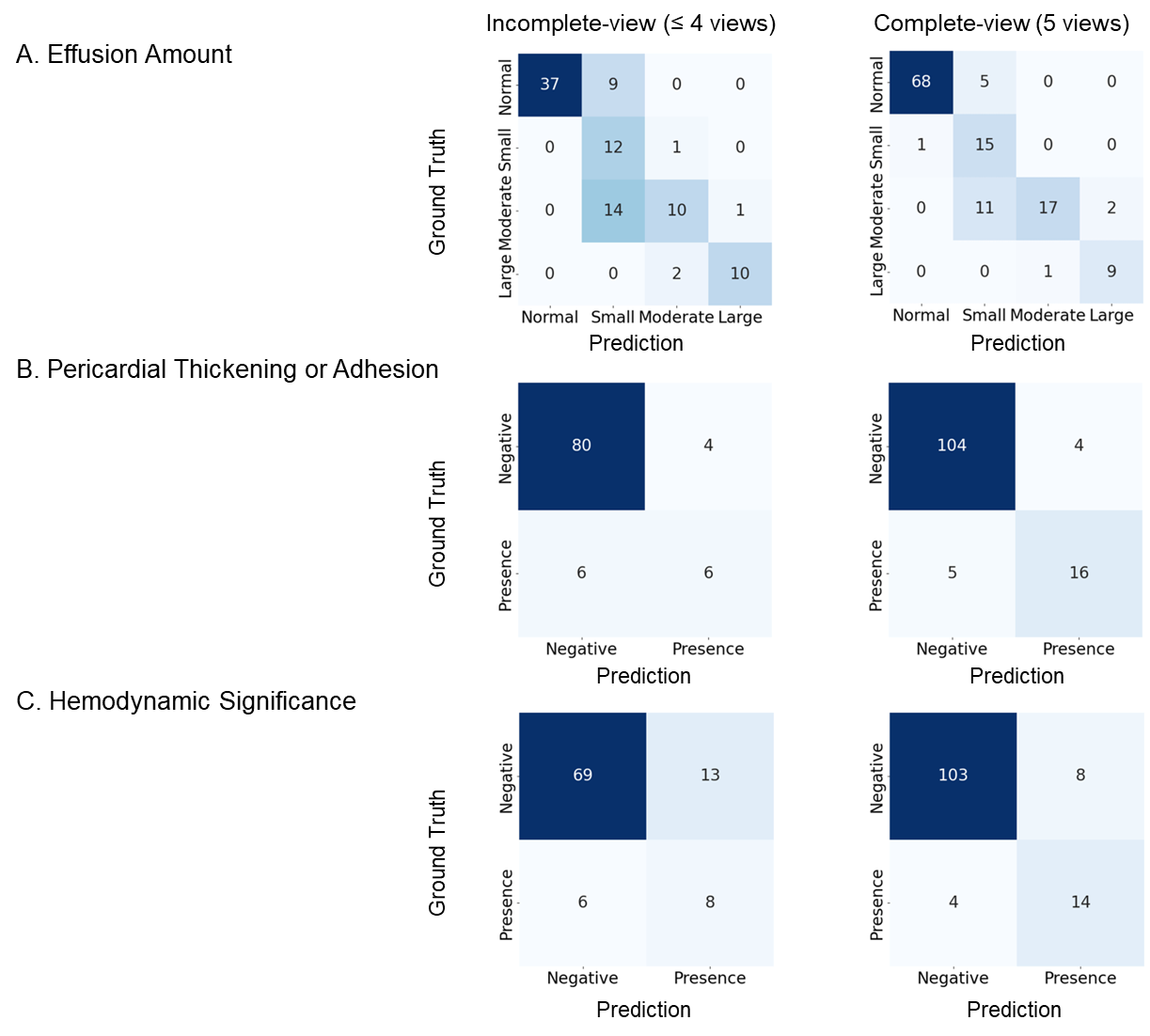

**4.2. Diagnostic Performance Metrices by View Completeness**

|  | **Incomplete view (≤ 4 views) (N=96)** | | | | |  | **Complete view (5 views) (N=129)** | | | | |
| --- | --- | --- | --- | --- | --- | --- | --- | --- | --- | --- | --- |
|  | **Accuracy** | **Precision** | **Sensitivity** | **Specificity** | **F1-score** |  | **Accuracy** | **Precision** | **Sensitivity** | **Specificity** | **F1-score** |
| **Pericardial Effusion** | | | | | | | | | | | |
| Normal | 0.906 | 1.000 | 0.804 | 1.000 | 0.892 |  | 0.954 | 0.986 | 0.932 | 0.982 | 0.958 |
| Small | 0.750 | 0.343 | 0.923 | 0.723 | 0.500 |  | 0.868 | 0.484 | 0.938 | 0.858 | 0.638 |
| Moderate | 0.813 | 0.769 | 0.400 | 0.958 | 0.526 |  | 0.892 | 0.944 | 0.567 | 0.990 | 0.708 |
| Large | 0.969 | 0.909 | 0.833 | 0.988 | 0.870 |  | 0.977 | 0.818 | 0.900 | 0.983 | 0.857 |
| **Pericardial Thickening/Adhesion** | | | | | | | | | | | |
| Presence | 0.896 | 0.600 | 0.500 | 0.952 | 0.545 |  | 0.930 | 0.800 | 0.762 | 0.963 | 0.780 |
| **Hemodynamic Significance** | | |  |  |  |  |  |  |  |  |  |
| Presence | 0.802 | 0.381 | 0.571 | 0.841 | 0.457 |  | 0.907 | 0.636 | 0.778 | 0.928 | 0.700 |

**4.3. ROC Curve Analysis by View Completeness**

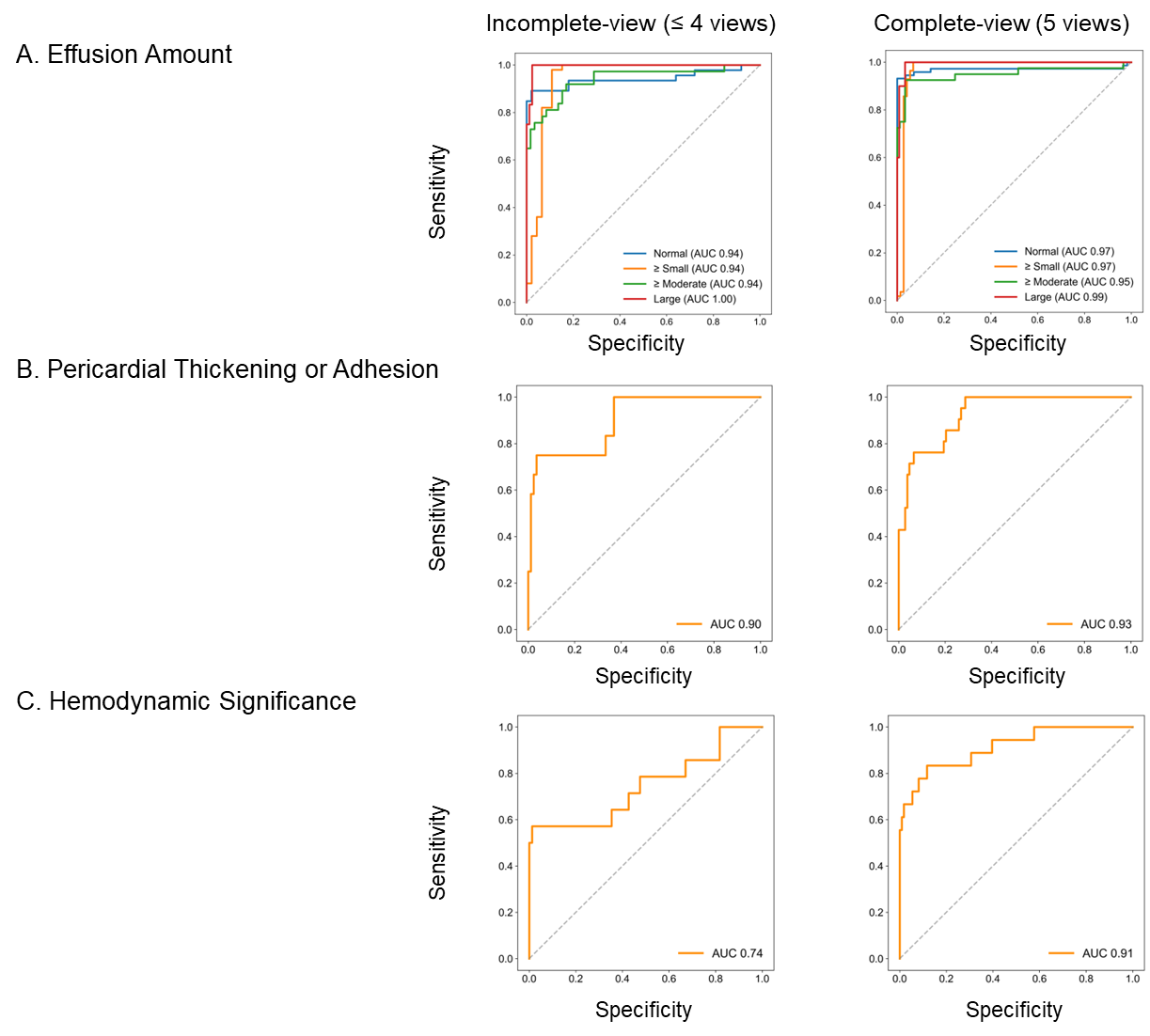

**Supplemental Results 5. Model Performance Stratified by Number of Available B-mode Video Clips (Upper Two Tertiles vs. Lower Tertile)**

**5.1. Confusion Matrices by Number of Available Video Clips**

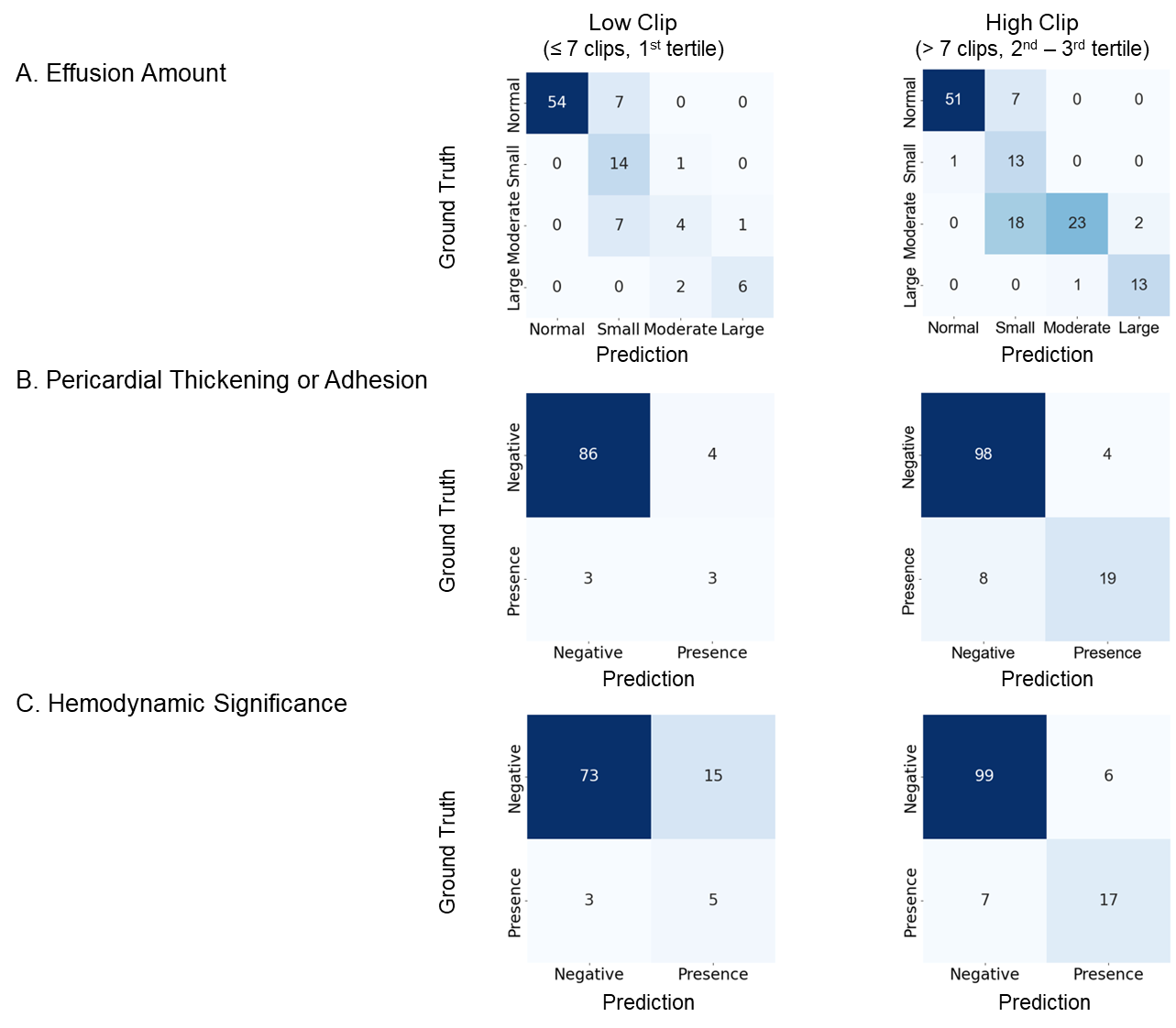

**5.2. Diagnostic Performance Metrics by Number of Available Video Clips**

|  | **Low-Clip (≤7 clips, 1st tertile) (N= 96)** | | | | |  | **High-Clip (> 7 clips, 2nd-3rd tertile) (N = 129)** | | | | |
| --- | --- | --- | --- | --- | --- | --- | --- | --- | --- | --- | --- |
|  | **Accuracy** | **Precision** | **Sensitivity** | **Specificity** | **F1-score** |  | **Accuracy** | **Precision** | **Sensitivity** | **Specificity** | **F1-score** |
| **Pericardial Effusion** | | | | | | | | | | | |
| Normal | 0.927 | 1.000 | 0.885 | 1.000 | 0.939 |  | 0.938 | 0.981 | 0.879 | 0.986 | 0.927 |
| Small | 0.844 | 0.500 | 0.933 | 0.827 | 0.651 |  | 0.798 | 0.342 | 0.929 | 0.783 | 0.500 |
| Moderate | 0.885 | 0.571 | 0.333 | 0.964 | 0.421 |  | 0.837 | 0.958 | 0.535 | 0.988 | 0.687 |
| Large | 0.969 | 0.857 | 0.750 | 0.989 | 0.800 |  | 0.977 | 0.867 | 0.929 | 0.983 | 0.897 |
| **Pericardial Thickening/Adhesion** | | | | | | | | | | | |
| Presence | 0.927 | 0.429 | 0.500 | 0.956 | 0.462 |  | 0.907 | 0.826 | 0.704 | 0.961 | 0.760 |
| **Hemodynamic Significance** | | |  |  |  |  |  |  |  |  |  |
| Presence | 0.812 | 0.250 | 0.625 | 0.830 | 0.357 |  | 0.899 | 0.739 | 0.708 | 0.943 | 0.723 |

**5.3. ROC Curve Analysis by Number of Available Video Clips**

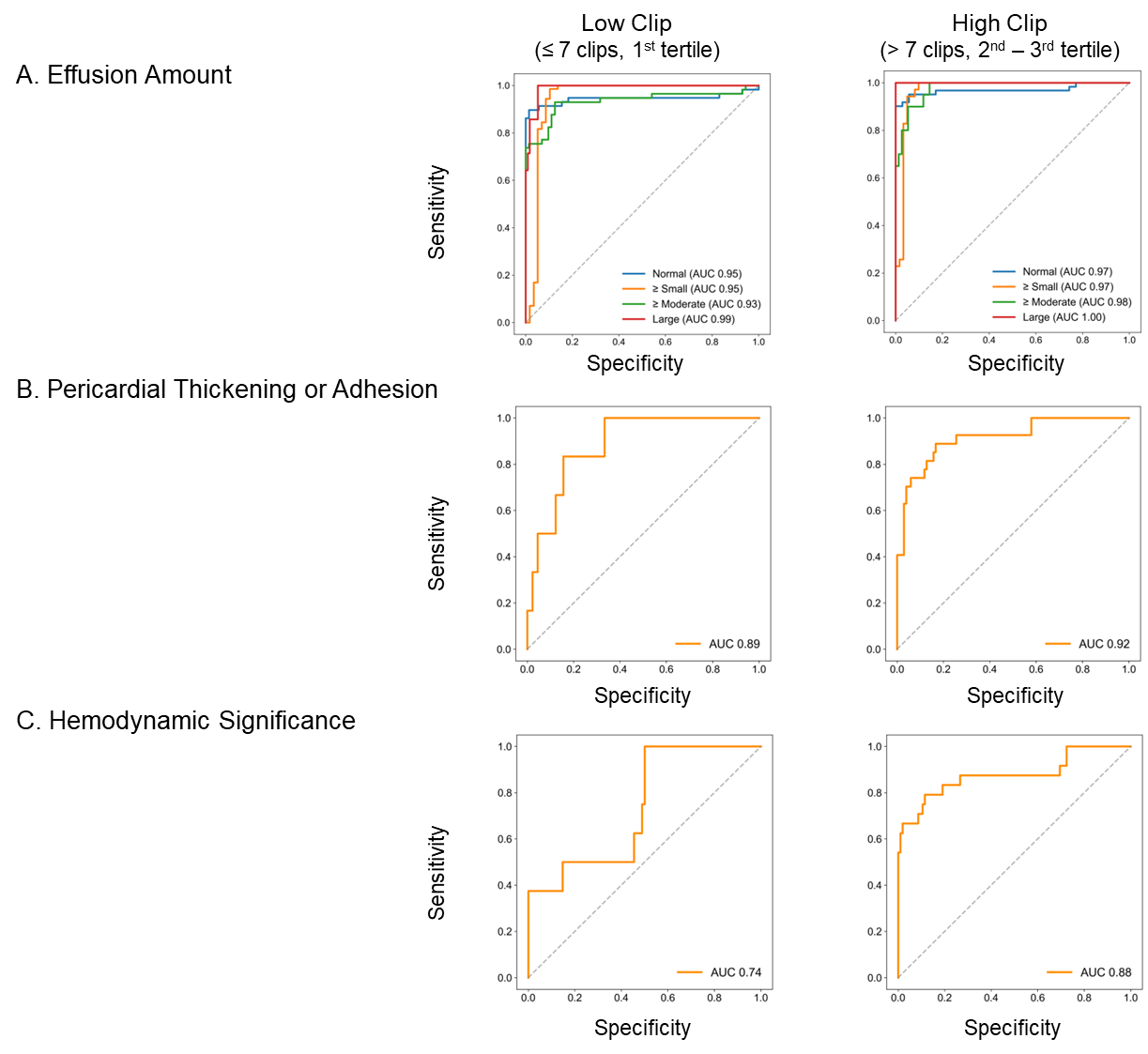
